# Micro RNA-based regulation of genomics and transcriptomics of inflammatory cytokines in COVID-19

**DOI:** 10.1101/2021.06.08.21258565

**Authors:** Manoj Khokhar, Sojit Tomo, Purvi Purohit

**Affiliations:** Department of Biochemistry, All India Institute of Medical Sciences, Jodhpur 342005, India

**Keywords:** Cytokine storm, immuno-interactomics, COVID-19, Cytokines, MicroRNA, SARS-CoV-2

## Abstract

**Background:** Coronavirus disease 2019 is characterized by the elevation of a wide spectrum of inflammatory mediators, which are associated with poor disease outcomes. We aimed at an in-silico analysis of regulatory microRNA and their transcription factors (TF) for these inflammatory genes that may help to devise potential therapeutic strategies in the future.

**Methods:** The cytokine regulating immune-expressed genes (CRIEG) was sorted from literature and the GEO microarray dataset. Their co-differentially expressed miRNA and transcription factors were predicted from publicly available databases. Enrichment analysis was done through mienturnet, MiEAA, Gene Ontology, and pathways predicted by KEGG and Reactome pathways. Finally, the functional and regulatory features were analyzed and visualized through Cytoscape.

**Results:** Sixteen CRIEG were observed to have a significant protein-protein interaction network. The ontological analysis revealed significantly enriched pathways for biological processes, molecular functions, and cellular components. The search performed in the MiRNA database yielded 10 (ten) miRNAs that are significantly involved in regulating these genes and their transcription factors.

**Conclusion:** An in-silico representation of a network involving miRNAs, CRIEGs, and TF which take part in the inflammatory response in COVID-19 has been elucidated. These regulatory factors may have potentially critical roles in the inflammatory response in COVID-19 and may be explored further to develop targeted therapeutic strategies and mechanistic validation.

## 1. Introduction

Cytokine storm in severe or critically ill coronavirus disease 2019 (COVID-19) patients is characterized by the elevation of a wide spectrum of inflammatory mediators. These include cytokines and chemokines, originating from airway epithelial cells as well as various immune cells, and act as independent risk factors for disease severity and mortality (Liu *et al*. 2020, p. 19).

Various cytokines and chemokines have been observed to play dominant roles in different stages of the COVID-19 disease. Association of COVID-19 severity and mortality with higher levels of interleukin-6 (IL-6) have been corroborated in various studies (Cummings *et al*. 2020, Hajifathalian *et al*. 2020, Ruan *et al*. 2020). However, depending upon the stage in the natural history of COVID-19 disease SARS CoV-2 has the menacing feature of longer persistence in the environment and various inanimate surfaces (Khokhar, Roy, *et al*. 2020) and different inflammatory mediators have been observed to play a dominant role in Acute kidney injury pathophysiology (Khokhar, Purohit, *et al*. 2020). Control this disease Newer diagnostic tools, based on the Clustered Regularly Interspaced Short Palindromic Repeats/Cas (CRISPR-Cas) system is used for better diagnostic accuracy.(Gadwal *et al*. 2021, p. 19) In the initial stage, when clinical symptoms are mild, the severe acute respiratory syndrome coronavirus 2 (SARS-CoV-2) replicates rapidly in blood (Chen, Lan, *et al*. 2020). Chemokines are the inflammatory mediators that characterize this initial stage. Chemokine (C-C motif) ligands (CCL), namely CCL8, CCL9, and CCL2 expression were found to be increased in the initial phase (Blanco-Melo *et al*. 2020, p. 19). In severe COVID-19 patients, serum CCL5 levels were elevated even before the occurrence of an IL-6 peak (Zhao *et al*. 2020). Further, in bronchoalveolar lavage (BAL) fluid of COVID-19 patients, a chemokine-rich signature was observed characterized by the expression of CCL2, CCL3, CCL4, CCL7, CCL8, chemokine (C-X-C motif) ligand 2 (CXCL2), CXCL8, CXCL17, and interferon-inducible protein 10 (IP-10) (Lu *et al*. 2020, Xiong *et al*. 2020).

During the amplification phase, inflammatory immune responses get aggravated and the disease progresses rapidly to severe/critical illness. The chemokines secreted in the initial phase recruit inflammatory innate and adaptive immune cells resulting in an exaggerated inflammatory immune response. Peripheral levels of inflammatory mediators including IL-2, IL-6, IL-7, IL-10, tumor necrosis factor-alpha (TNF-α), CCL-2, and CCL-3 were highly elevated in this phase (Chen, Wu, *et al*. 2020, Huang *et al*. 2020). The unchecked elevation of inflammatory mediators leads to vascular leakage, complement cascade activation, and cytokine storm in this consummation phase (Wang *et al*. 2020, Yang *et al*. 2020, Zhou *et al*. 2020). Apart from the inflammatory mediators, it had also been observed that the mRNA expression levels of inflammatory genes peaked as the respiratory function deteriorated (Ong *et al*. 2020). Further, COVID-19 patients had also shown the possibility of alterations in transcription factors, affecting both cytokines as well as immune cells, adding another layer of dimension to the regulation and release of cytokines in a cytokine storm (Claverie 2020, p. 19, De Biasi *et al*. 2020).

This study aimed to search the various databases for miRNAs that can affect the genetic expression of these inflammatory mediators and the transcription factors that regulate the expression. The insight into the regulation of the expression of these cytokine genes by miRNAs and transcription factors will help in devising better targeted therapies to address the complications in severe COVID-19 disease due to cytokine storm.

## 2. Methodology

### 2.1 Identification of cytokine responsible for inflammation and cytokine storm in SARS-CoV-2

Several keywords including “Inflammation”, “Immunity”, “Immunogenetics”, “Cytokine storm”, “Acute respiratory distress syndrome”, “ARDS”, “COVID-19”, “cytokines”, “Coronavirus disease”, “SARS-CoV-2” and “Severe Acute Respiratory Syndrome” and “19990101 to 20200706” were searched in PubMed **(Figure 1, Supplementary Tables 1 & Table-1A)**.

**Figure 1.**
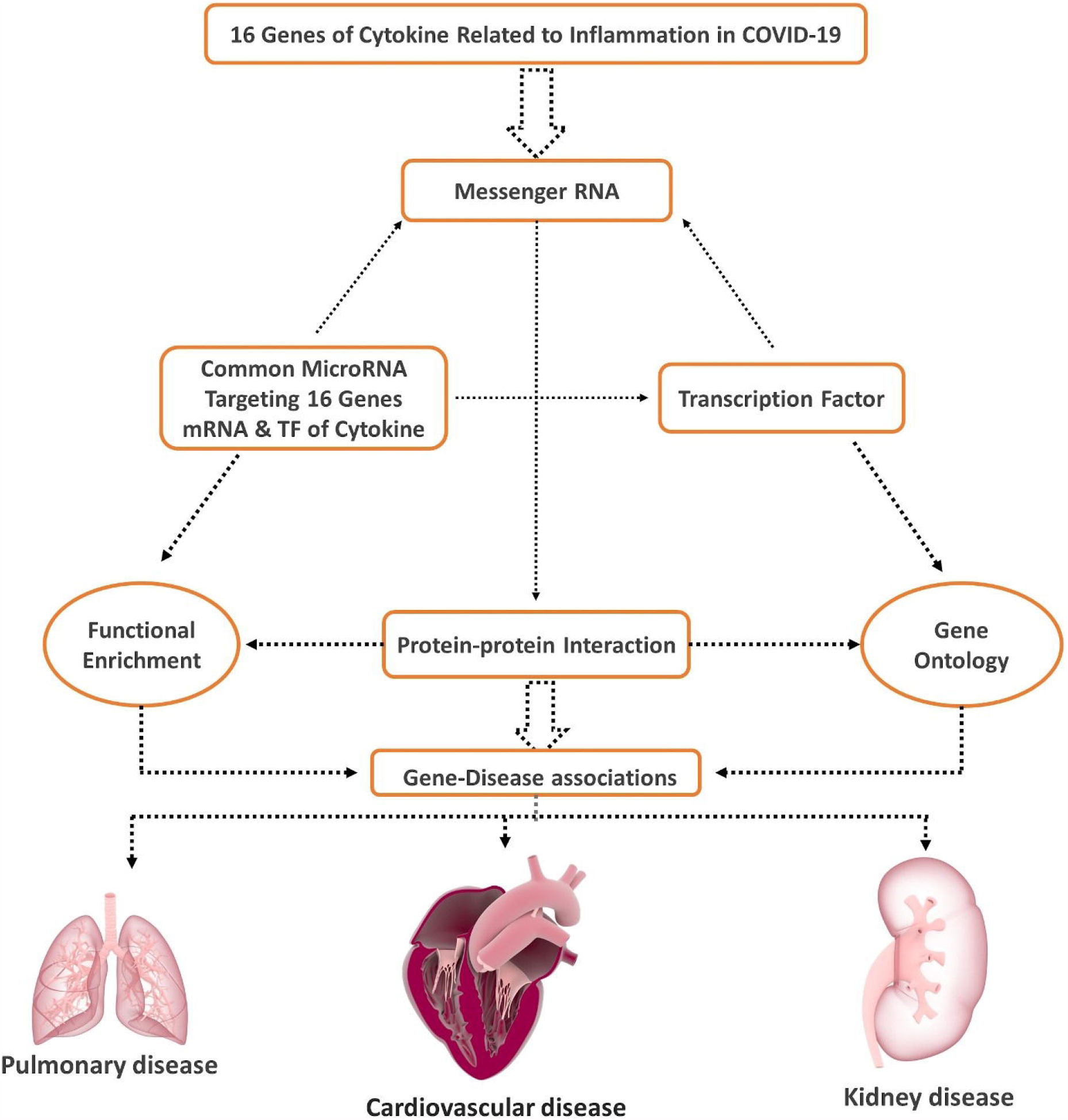
Flow Chart of the data processing and Analysis.

**Figure 1B.**
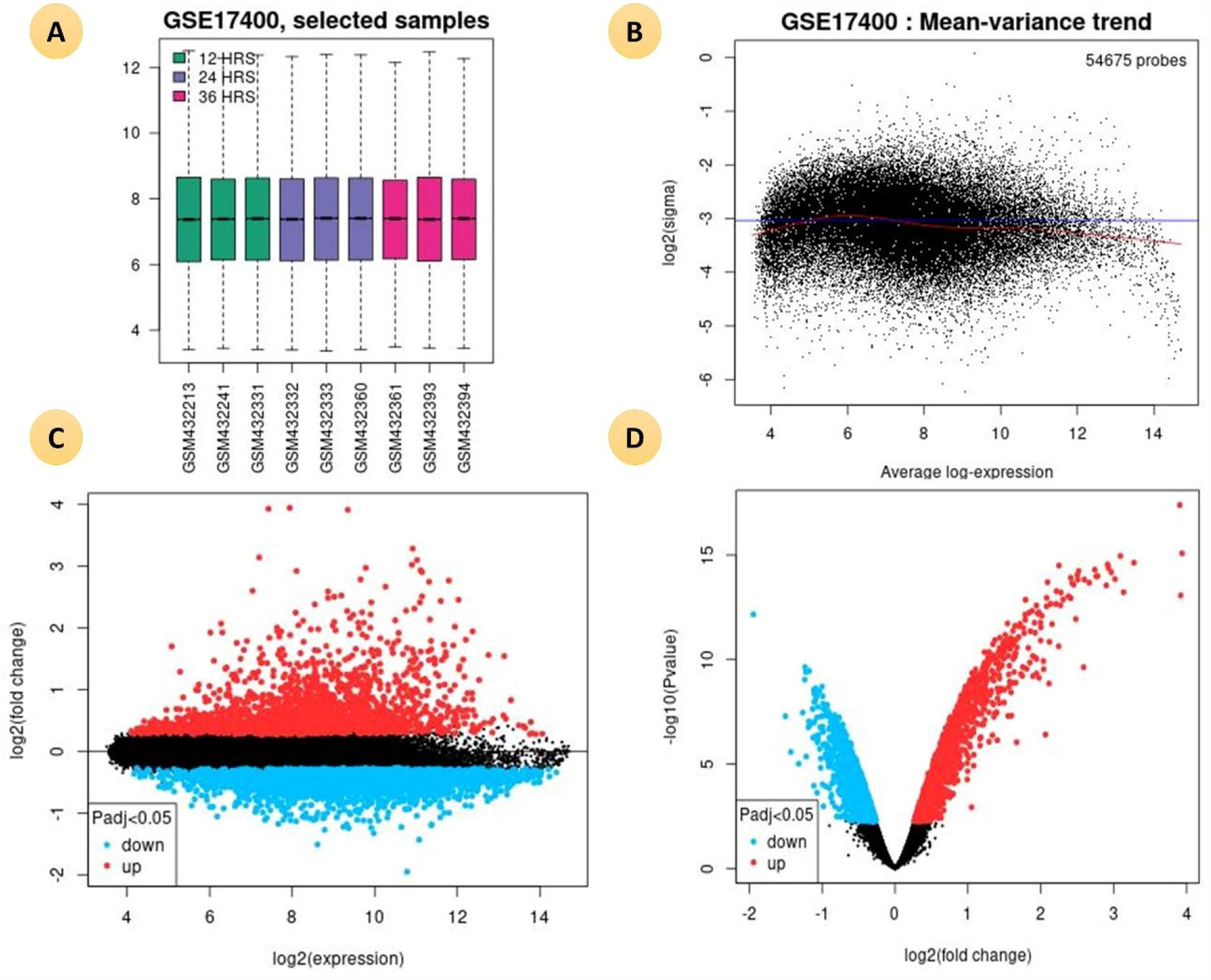
(A). Boxplot is represent the total DEGs of diffrent groups for study samples. (B). Mean-variance trend plot is applicable to check the mean-variance relationship of the DEGs data, after fitting a linear model. (C). A mean difference (MD) plot displays log2 fold change versus average log2 expression values of DEGs. (D) A volcano plot displays statistical significance (-log10 P value) versus magnitude of change (log2 fold change) DEGs.

**Table: 1 A.** List of Cytokine Strom genes and there regulating transcription factors expressed in GSE17400.

| Cytokine Strom Gene |  |  | Transcription Factors of Cytokine Gene |  |  |
| --- | --- | --- | --- | --- | --- |
| Gene Symbol | P Value | Fold Change | Gene Symbol | P Value | Fold Change |
| CCL2 | 0.01 | 6.07 | AHR | 0.03 | 4.25 |
| CCL4 | 0.03 | 4.59 | CREM | 0.05 | 3.75 |
| CXCL10 | 0.00 | 98.60 | DDIT3 | 0.00 | 52.60 |
| CXCL8 | 0.00 | 56.40 | E2F1 | 0.00 | 10.50 |
| IL6 | 0.00 | 139.00 | EGR1 | 0.00 | 75.60 |
| IL7 | 0.00 | 24.60 | EP300 | 0.01 | 6.13 |
| JAK2 | 0.00 | 16.40 | ESR1 | 0.00 | 7.64 |
| TNF | 0.02 | 5.26 | ETS2 | 0.00 | 82.10 |
|  |  |  | HDAC1 | 0.03 | 4.49 |
|  |  |  | HDAC2 | 0.00 | 9.29 |
|  |  |  | IRF1 | 0.00 | 132.00 |
|  |  |  | JUN | 0.00 | 45.20 |
|  |  |  | KLF4 | 0.00 | 58.10 |
|  |  |  | NFAT5 | 0.03 | 4.46 |
|  |  |  | NFKB1 | 0.00 | 12.70 |
|  |  |  | NFKBIA | 0.00 | 81.60 |
|  |  |  | REL | 0.00 | 14.20 |
|  |  |  | RUNX1 | 0.04 | 4.10 |
|  |  |  | SIRT1 | 0.00 | 26.40 |
|  |  |  | SP100 | 0.00 | 111.00 |
|  |  |  | SP140L | 0.00 | 53.80 |
|  |  |  | STAT1 | 0.00 | 80.60 |
|  |  |  | XBP1 | 0.02 | 4.81 |
|  |  |  | ZFP36 | 0.00 | 14.40 |

### 2.2 Microarray Data collection

We have searched in GEO database by several keywords including “SARS”, “Corona Virus”, “Blood”, “Homo sapiens”, “Expression profiling by array”, “bronchial epithelial cells” from 01/01/2012 to 17/12/2020. Selected one gene series expressions (GSEs) data were for further study. GSE17400 contain bronchial epithelial cells of 09 (nine) samples. **(Table-1B)**.

### 2.3 Identification of co-differentially expressed mRNAs *responsible for inflammation and cytokine storm in SARS-CoV*

GEO2R is an online interactive web tool used to compare two or more groups of samples in a GEO Series to identify genes that are differentially expressed across experimental conditions. We obtain differentially expressed genes (DEGs) from two datasets (GSE17400) for innate immune responses of human bronchial epithelial cells against SARS-CoV with the help of GEO2R with the cutoff criteria of p<0.05. Common genes in both datasets were identified and isolated with the use of the Venn-diagram. (*Figure 1*)

### 2.4 Identification of common transcriptome related to cytokine regulating immune expressed genes (CRIEGs)

Cytokines are mostly regulated at the transcriptional level by specific combinations of TFs that recruit cofactors and the transcriptional machinery (Carrasco Pro *et al*. 2018). We identified the common transcription factors of CRIEGs through five different databases-TRRUST, RegNetwork, ENCODE, JASPAR, and CHEA. The targeted transcription factor of 16 cytokines were identified by well-established TF - target prediction database miRNet Version 2 (a miRNA-centric network visual analytics platform) (Chang *et al*. 2020). A co-regulatory transcriptome network was created based on inter-correlation in Cytoscape software. **(Table 2)**

**Table: 2B.** List of Cytokine Strom genes, transcription factors and common targeting microRNAs.

| Cytokine Gene | Transcription Factor | MicroRNAs |
| --- | --- | --- |
| IL1B | AHR | hsa-miR-106a-5p |
| IL2 | CEBPA | hsa-miR-155-5p |
| IL7 | CREM | hsa-miR-98-5p |
| CXCL8 | DDIT3 | hsa-miR-24-3p |
| IL9 | E2F1 | hsa-miR-204-5p |
| IL10 | EGR1 | hsa-miR-124-3p |
| IL17A | EP300 | hsa-miR-203a-3p |
| CSF3 | ESR1 | hsa-miR-335-5p |
| CSF2 | ETS2 | hsa-let-7c-5p |
| JAK2 | FOXP3 | hsa-miR-1-3p |
| TNF | HDAC1 |  |
| CXCL10 | HDAC2 |  |
| CCL2 | HSF1 |  |
| CCL3 | IRF1 |  |
| CCL4 | JUN |  |
| IL6 | KLF4 |  |
|  | NFAT5 |  |
|  | NFKB1 |  |
|  | NFKBIA |  |
|  | NR1I2 |  |
|  | REL |  |
|  | RELA |  |
|  | RUNX1 |  |
|  | SIRT1 |  |
|  | SP1 |  |
|  | SPI1 |  |
|  | STAT1 |  |
|  | STAT3 |  |
|  | VDR |  |
|  | XBP1 |  |
|  | ZFP36 |  |
|  | ZNF300 |  |

**Table 3:**
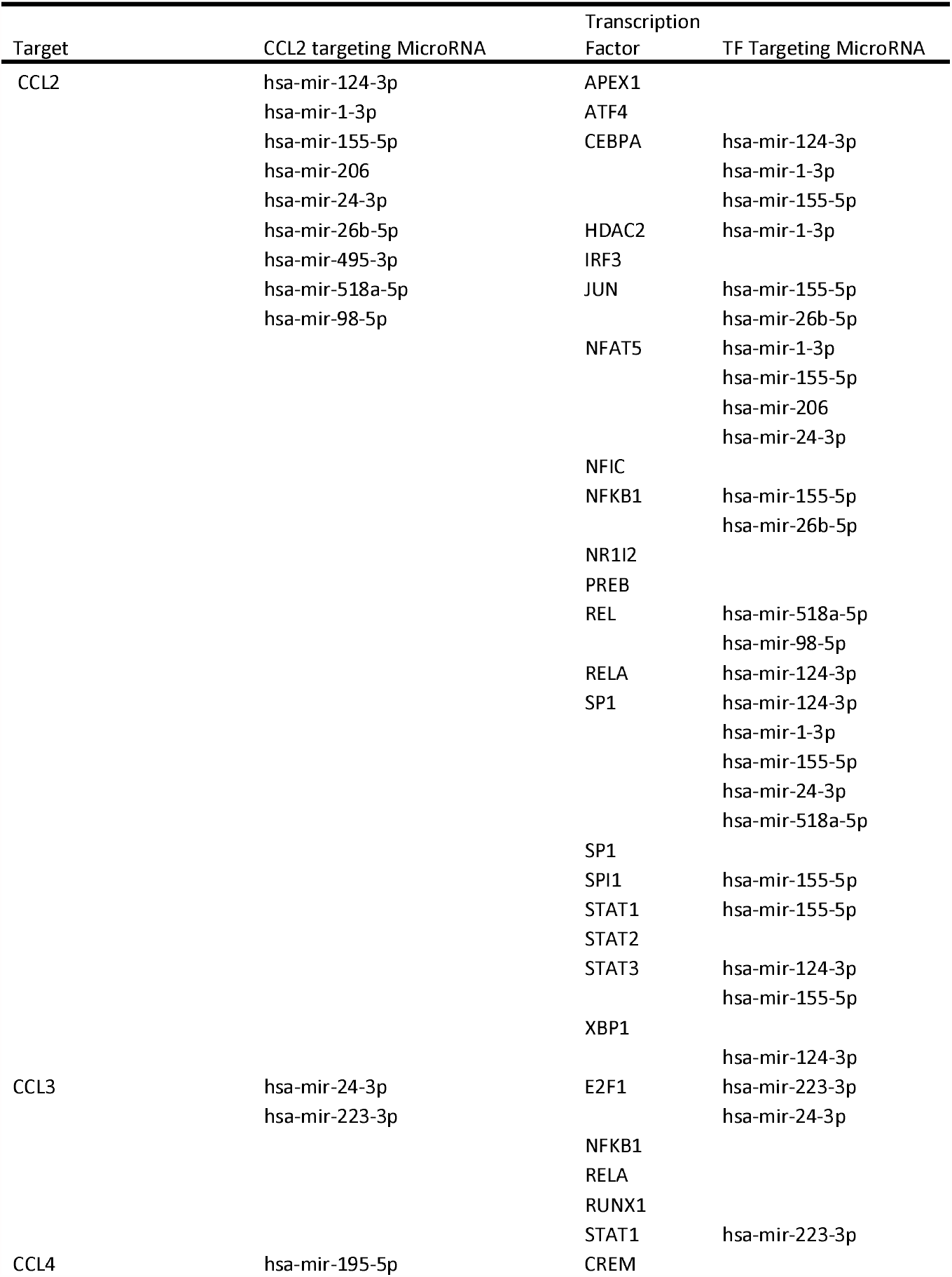
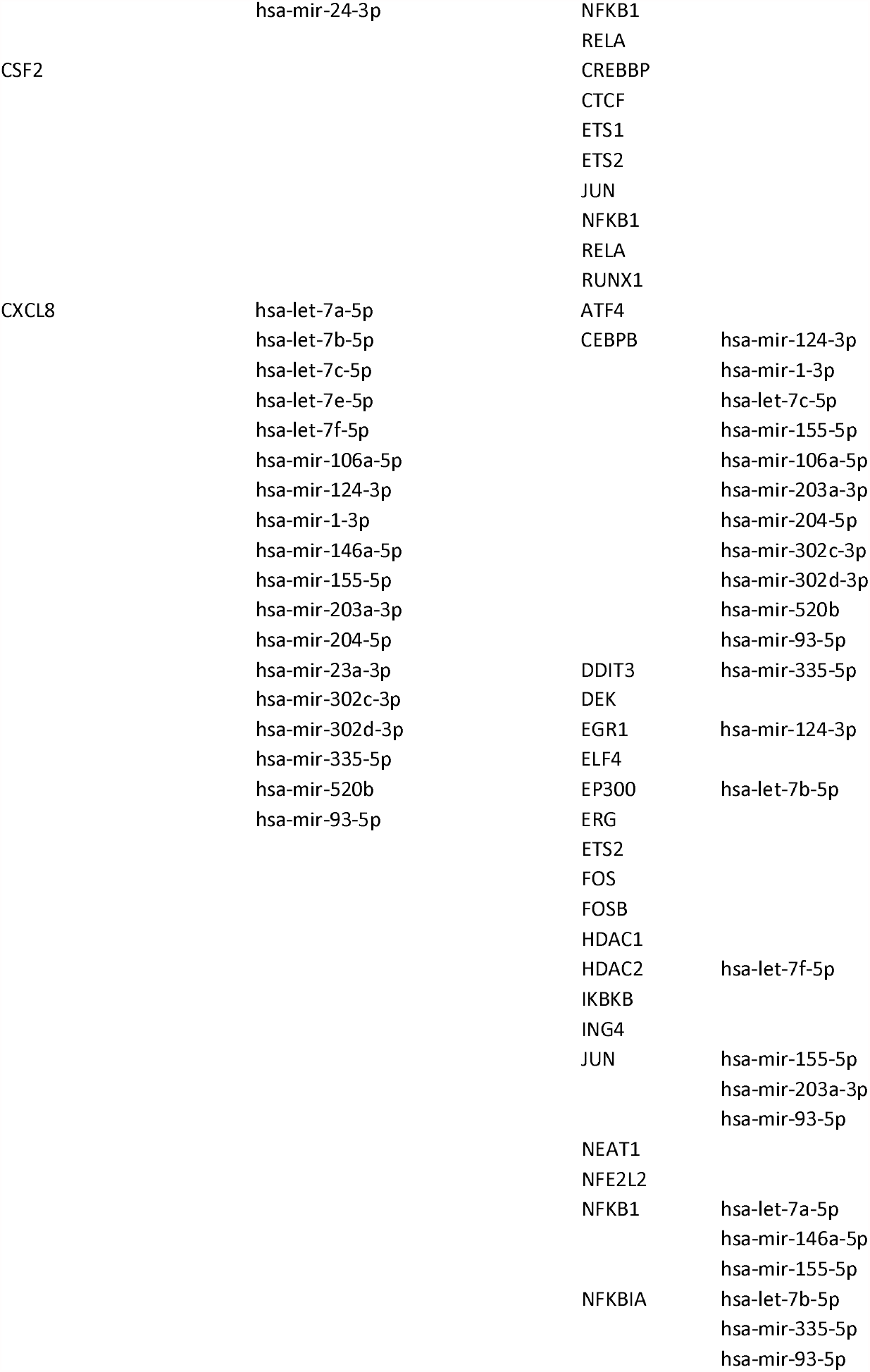
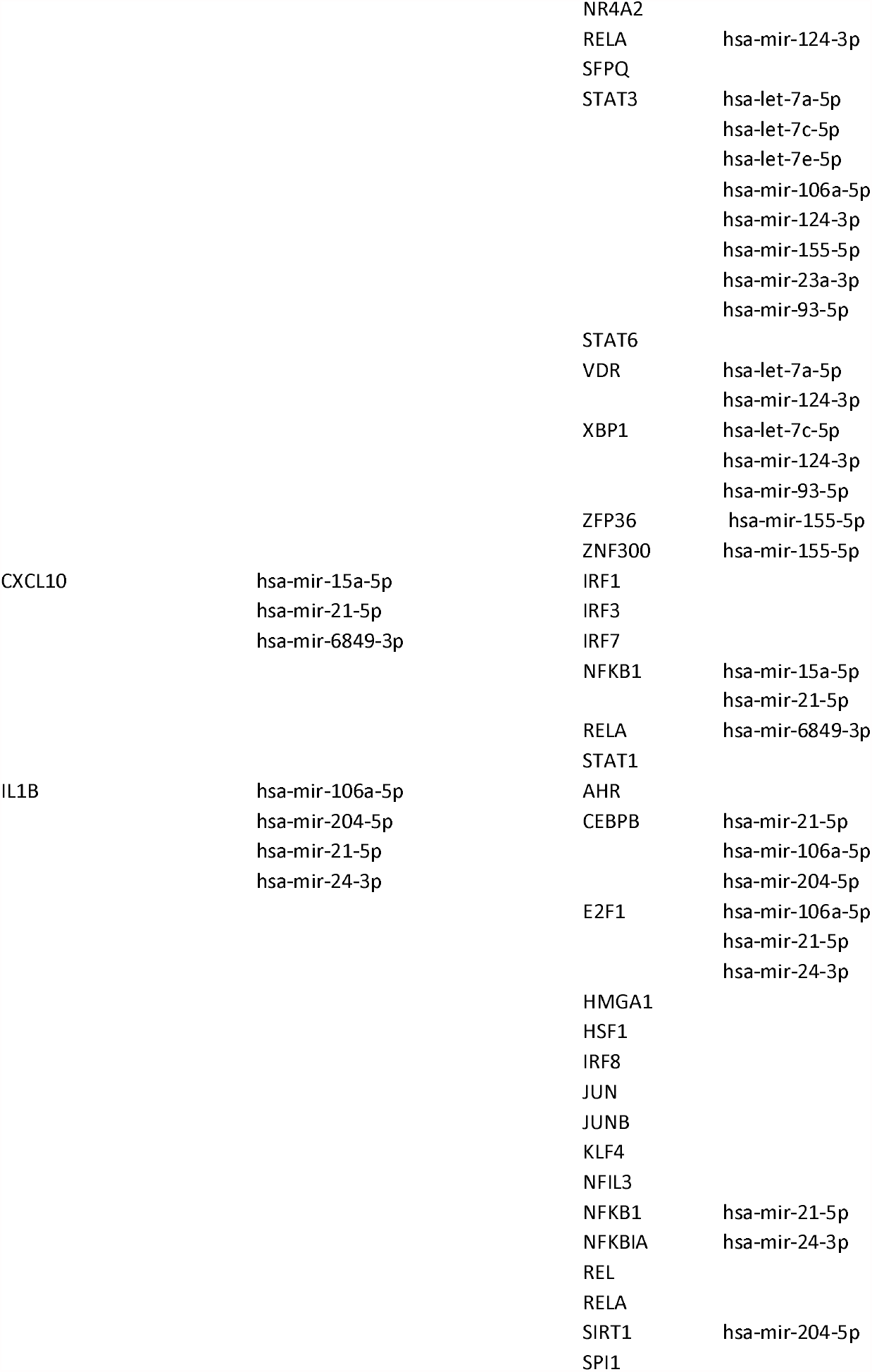
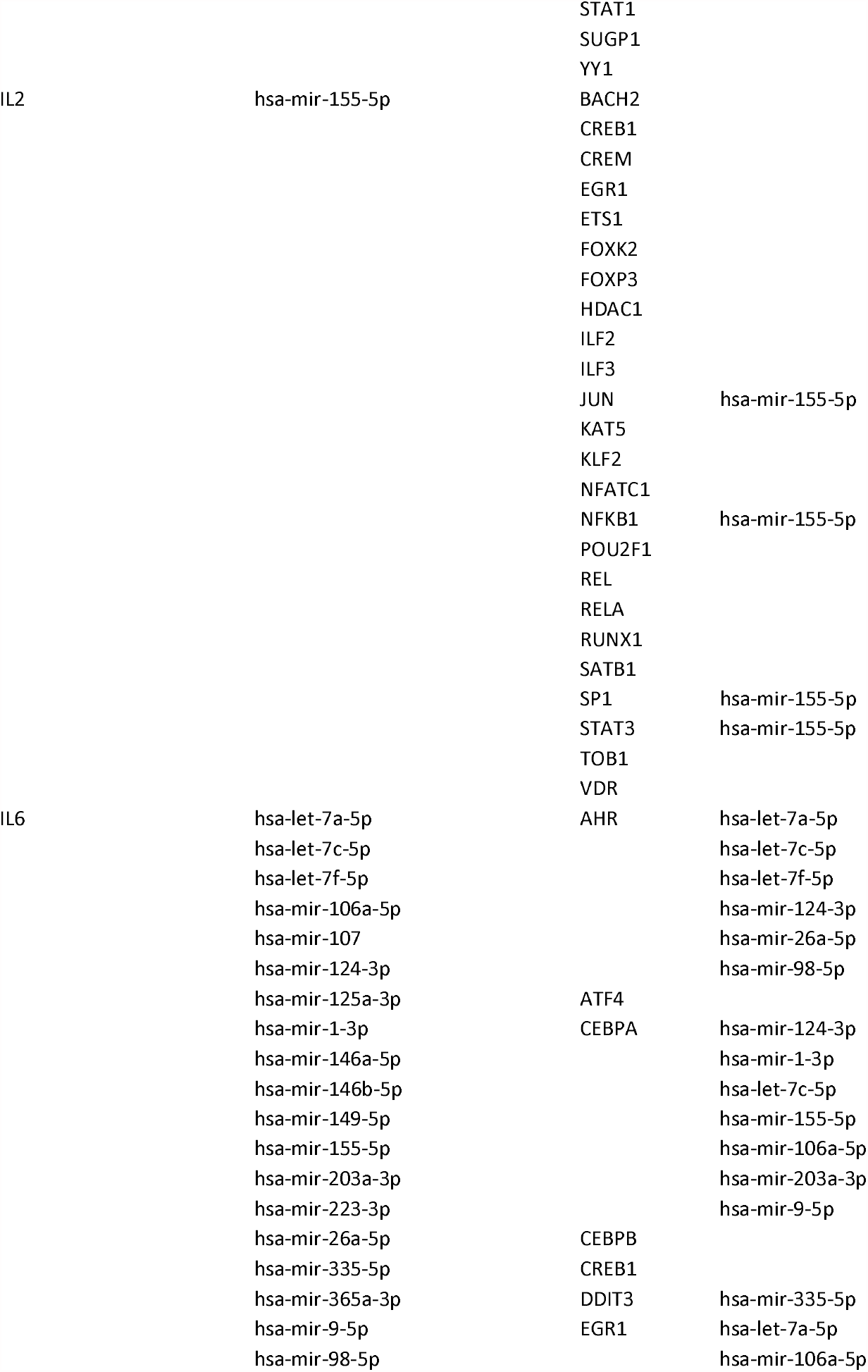
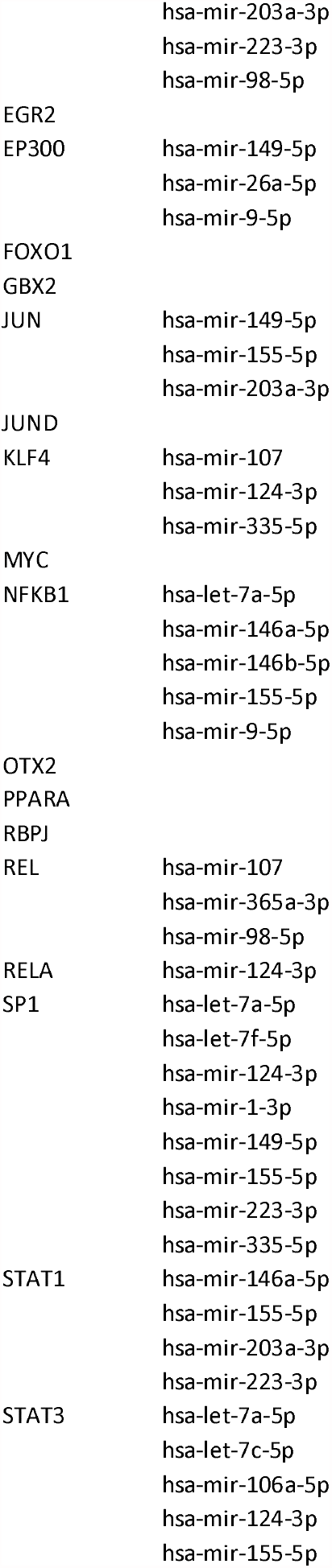
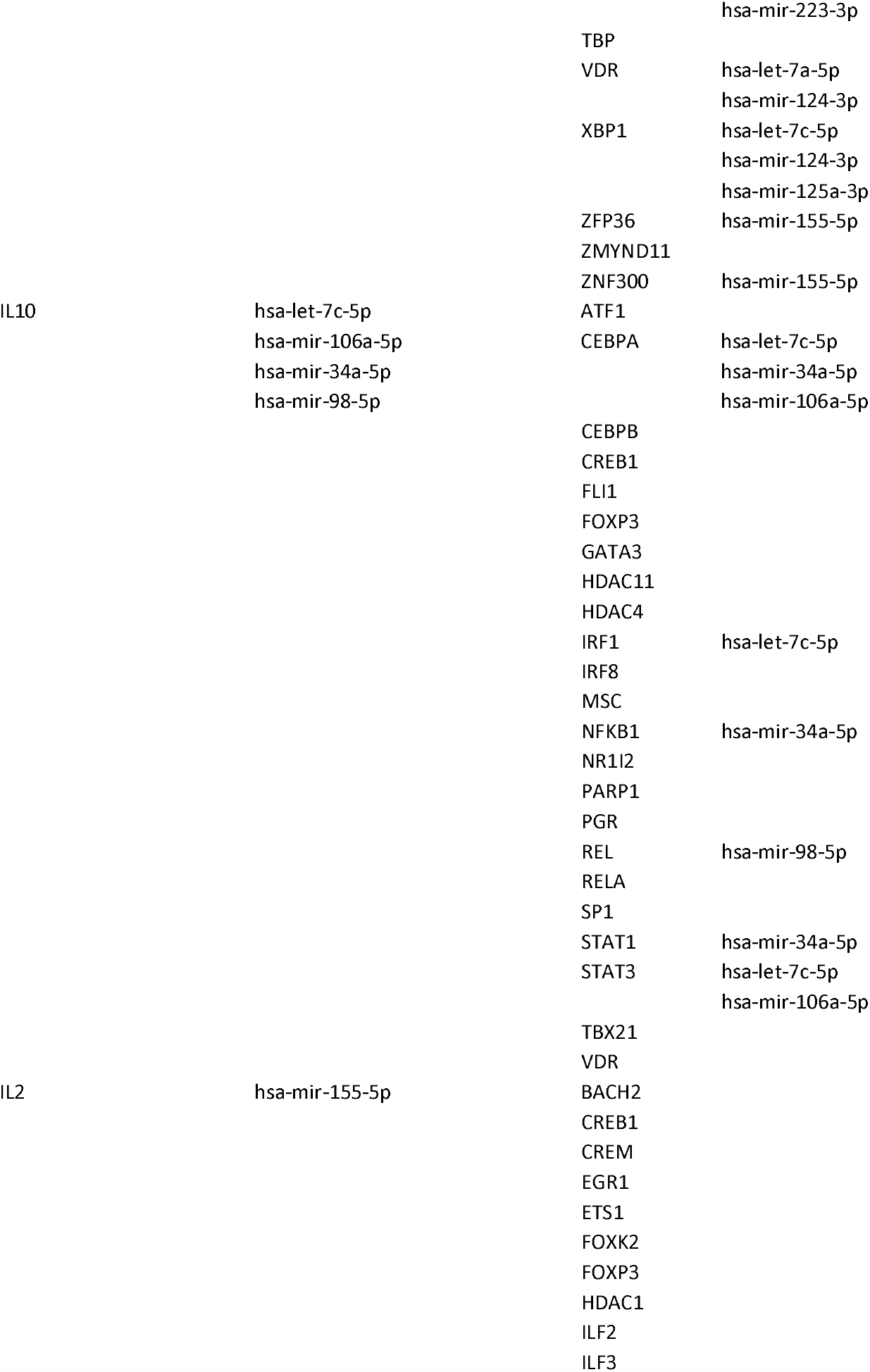
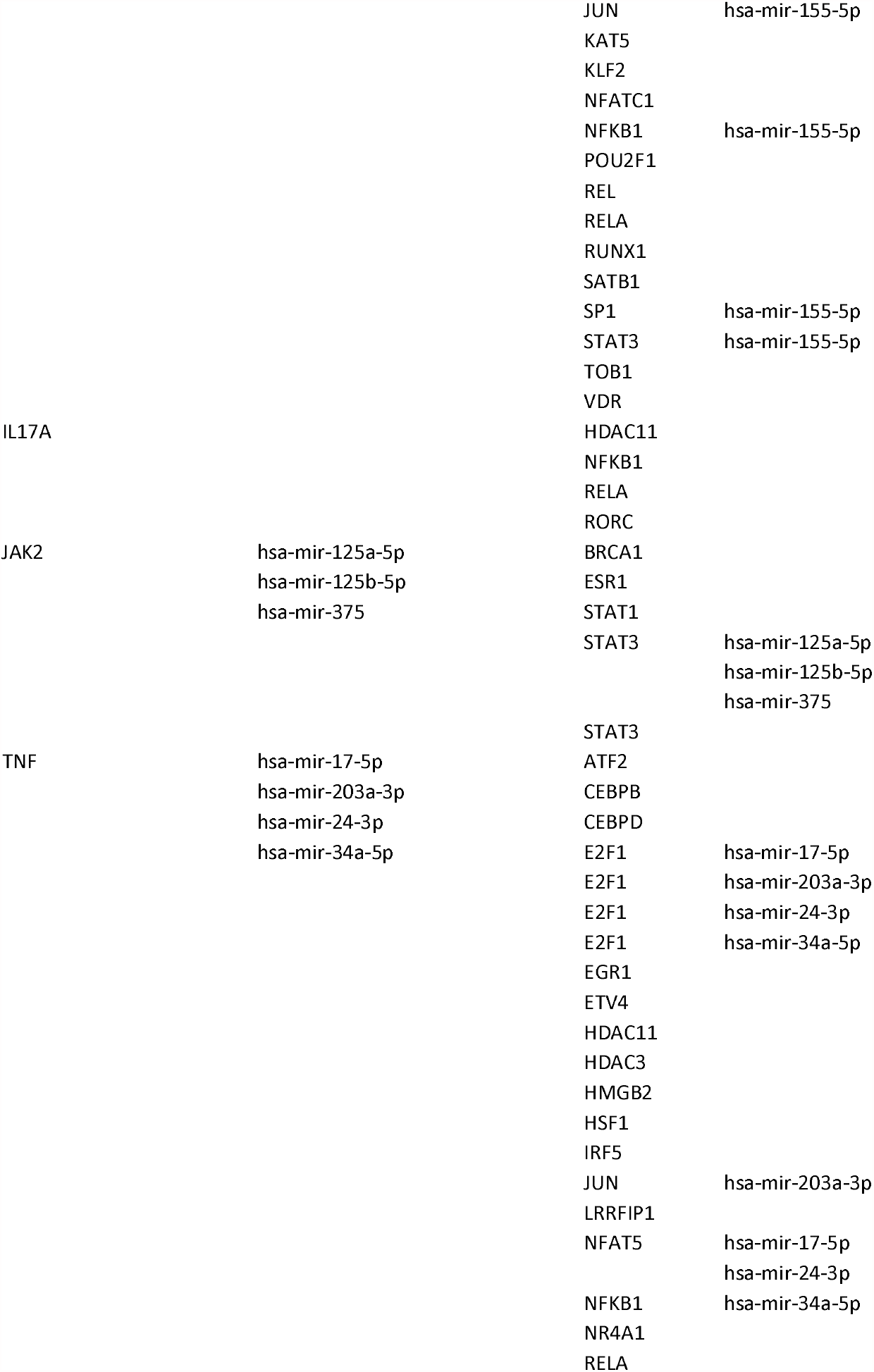
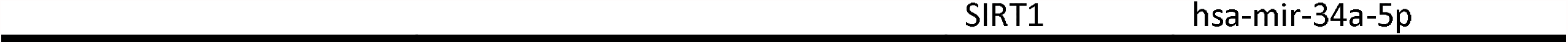
Cytokine and regulating transcription factor and common targeting MiroRNA of Cytokine and Its Transcription factor.

### 2.5 Identification and assortment of co-differentially regulated miRNAs of CRIEGs and transcriptome of CRIEGs

Evolutionary conserved small non-coding RNA or MicroRNA affect the gene expression by binding to specific mRNAs and regulate cell growth, differentiation, and death. miRNAs regulate multiple functions of T-cell subsets through immune homeostasis and immune tolerance that control the development, survival and activation (Garavelli *et al*. 2018). The targeted miRNA of cytokine and its transcriptome genes were predicted by well-established different miRNA target prediction databases miRDB, miRBase, miRNet Version 2 and TargetScan (Chang *et al*. 2020). A co-expressed network was created based on regulatory correlation analysis of Cytoscape software. **(Tables 1 & 2)**

### 2.6 Protein-protein interaction, functional enrichment and KEGG pathway analysis of CRIEGs and transcriptome of CRIEGs

The Search Tool for the Retrieval of Interacting Genes/Protein [STRING] (http://string-db.org/) was used to construct a protein-protein interaction (PPI) network using only overlapped DEGs and greater than 0.4 confidence score cut-off. The interaction networks for all 16 cytokines were constructed by Cytoscape (Shannon 2003, Otasek *et al*. 2019).

Analysis of the functional and regulatory features was carried out through gene ontology (GO), KEGG pathways through DAVID (the database for annotation, visualization and integrated discovery) and STRING (functional protein association networks) biological databases. **(Supplementary Table 2)**

### 2.7 Functional enrichment, disease relationship, and KEGG pathway analysis of co-differentially regulated miRNAs of CRIEGs and transcriptome of CRIEGs

Cytokine regulating immune expressed genes (CRIEGs) and transcriptome of CRIEGs are regulated by ten (10) common microRNAs. We will identify the all ten microRNAs enrichment of two database MIENTURNET and MiEAA (Licursi *et al*. 2019, Kern *et al*. 2020). MIENTURNET and miEAA (miRNA Enrichment Analysis and Annotation tool) perform both statistical and network-based analyses pathways and disease-related activity in cellular processes.

## 3. Results

### 3.1 Identification of - Co Expressed DEGs responsible for inflammation and cytokine storm in SARS-CoV

We searched for genes that were obtained from the literature search and their transcription factors that control them in the SARS-CoV dataset (GSE17400), most of which were expressed here. **(Figure 1B)**.

### 3.2 Identification of cytokine regulating immune expressed genes and construction of PPI Network

We found 16 cytokine regulating immune expressed genes (*IL-1b, IL-2, IL-7, IL-8, IL-9, IL-10, IL-17, G-CSF, GM-CSF, IFN-γ, TNF-α, CXCL10, MCP1, MIP1A, MIP1B, and IL-6*) from literature which is responsible for acute respiratory distress syndrome (ARDS) in COVID-19. Over-expression of these genes in a short time increases the severity of the disease. All 16 CRIEGs show interactions among themselves, based on the STRING database **(Figure 2B)**. The PPI network consisted of 16 nodes and 117 edges, the average local clustering coefficient was 0.977, and PPI enrichment p-value was highly significant (p<0.001).

**Figure 2A.**
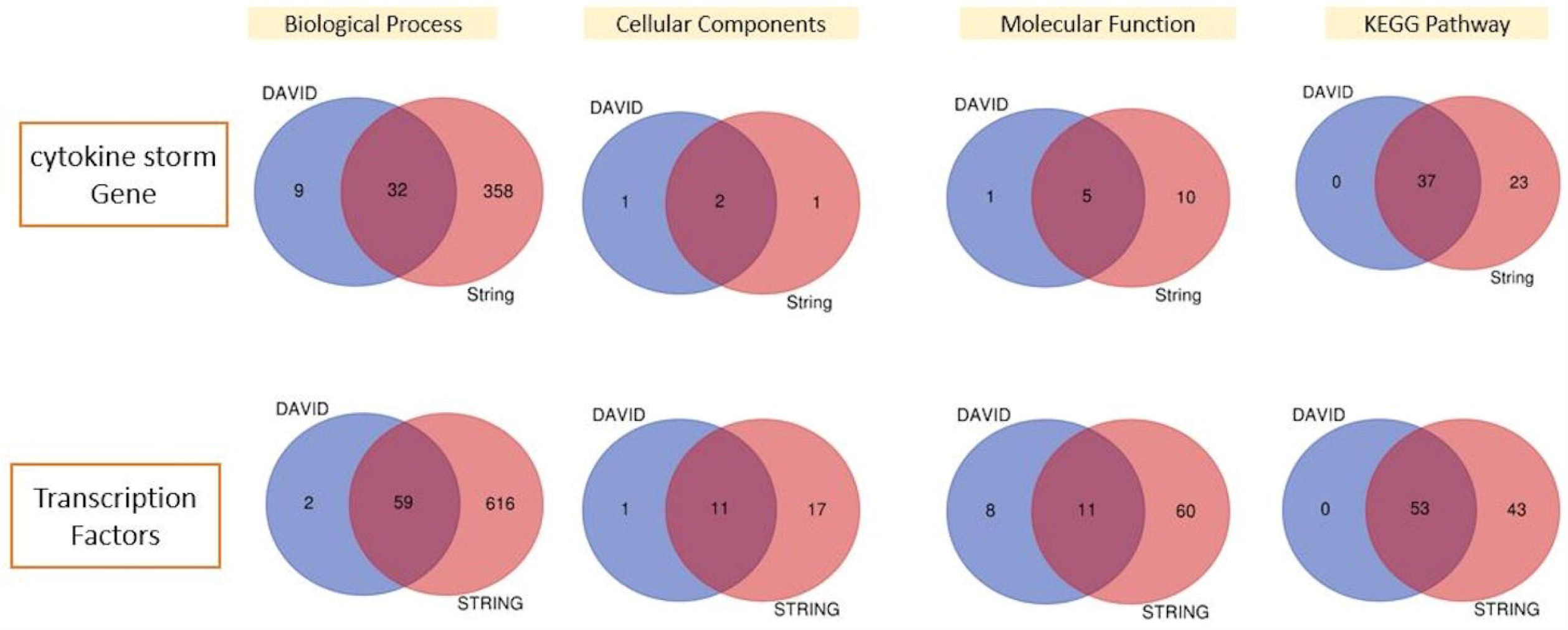
Venn diagram of enriched gene ontology

**Figure 2B.**
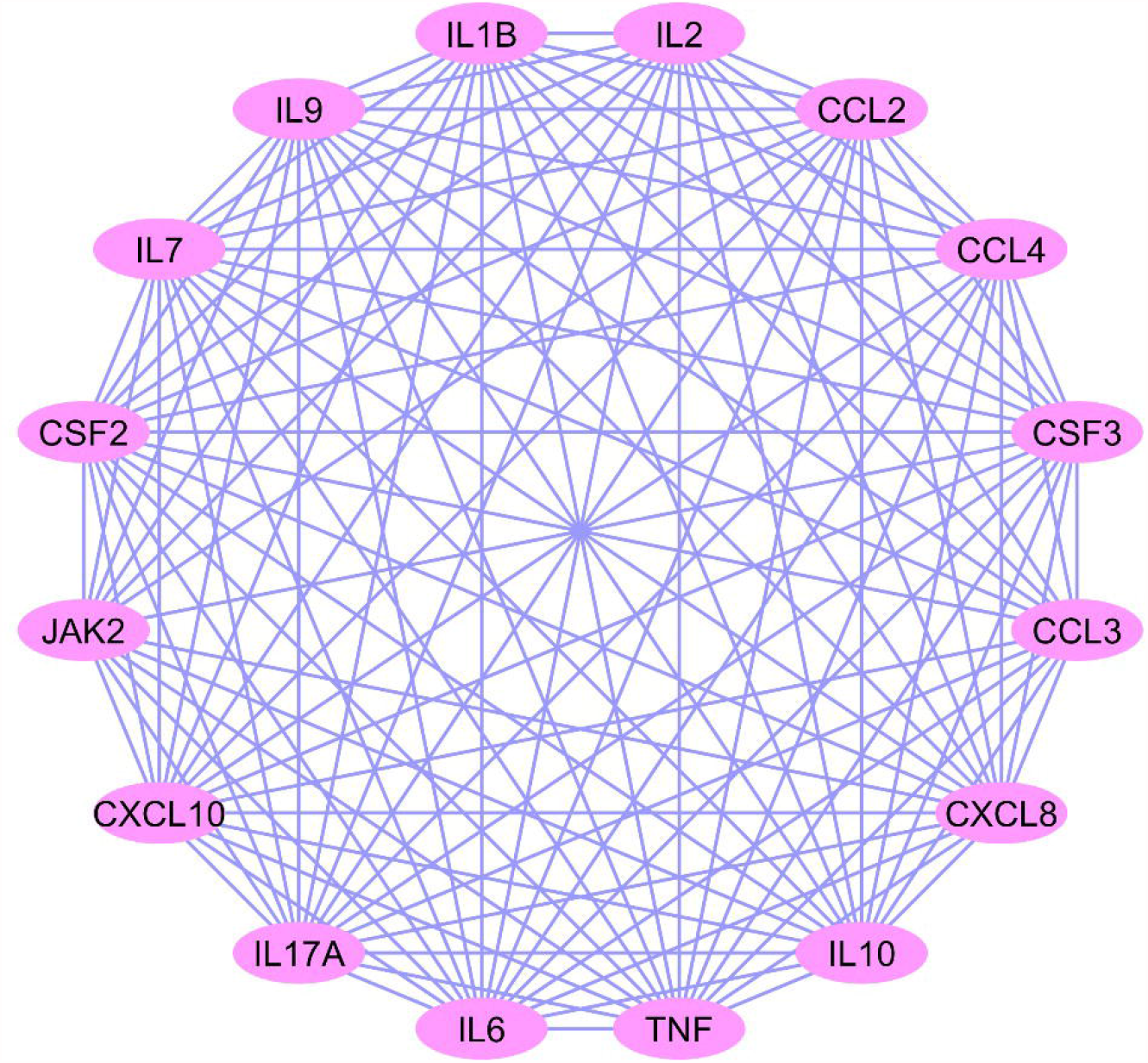
Protein-Protein Interaction between Cytokine storm genes.

### 3.3 Ontological analysis of CRIEGs

To get insights into the biomolecular significance of the identified CRIEGs, we performed gene ontology analysis by various databases and obtained enriched GO terms. STRING and DAVID were used to conduct the gene ontology analysis for CRIEGs within three different categories like biological process, molecular function and cellular component. Common statistically significant (p<0.05) ontological processes have been identified through DAVID.

Molecular process like Chemokine, Cytokine and Growth factor activity, Granulocyte colony-stimulating factor receptor binding, CC chemokine receptor (CCR), and C-X-C chemokine receptor (CXCR) binding play significant roles. Some crucial biological process such as immune and inflammatory response, chemokine-mediated signaling pathway, negative regulation of extrinsic apoptotic signaling pathway in absence of ligand, protein kinase B signaling, cellular response to interleukin-1, chemotaxis, receptor biosynthetic process, positive regulation of interleukin-23 production, estradiol secretion, myeloid cell differentiation, podosome assembly, osteoclast differentiation, tyrosine phosphorylation of Stat5 protein, cytokine secretion involved in immune response, cytokine secretion, transcription from RNA polymerase II promoter, lymphocyte, monocyte, neutrophil chemotaxis, cell proliferation, B cell proliferation, interferon-gamma production, inflammatory response response to glucocorticoid, regulation of cell proliferation, interleukin-6 production, negative regulation of growth of symbiont in host lipopolysaccharide-mediated signaling pathway, negative regulation of myoblast differentiation are regulated by CRIEGs. Cellular components (CC) found very limited organelles of the cell-like external side of the plasma membrane, extracellular region and extracellular space **(Figure 2 & Supplimentry Tables 3A, 3C and 3D)**.

### 3.4 Common Pathway enrichment analysis CRIEGs

STRING and DAVID were assessed to acquire KEGG pathways enriched by CRIEGs. Both databases were selected for preferred and significant (p<0.01) common pathways. Important selected pathways are enlisted in **(Supplimentry Table 3B)**.

### 3.5 Identification of common transcription factors of Cytokine Genes

We identified the common transcription factors of CRIEGs through TRRUST, RegNetwork, ENCODE, JASPAR and CHEA databases. During this identification process, we found a total of 32 transcription regulators. All transcription regulators *AHR, CEBPA, CREM, DDIT3, E2F1, EGR1, EP300, ESR1, ETS2, FOXP3, HDAC1, HDAC2, HSF1, IRF1, JUN, KLF4, NFAT5, NFKB1, NFKBIA, NR1I2, REL, RELA, RUNX1, SIRT1, SP1, SPI1, STAT1, STAT3, VDR, XBP1, ZFP36, ZNF300* commonly regulated the transcription of 16 CRIEGs **(Tables 1 & 2)**.

### 3.6 Ontological analysis of common transcription factors

To get deep insights into the biomolecular significance of the identified common TFs. We performed gene ontology analysis by various databases and obtained enriched GO terms. STRING and DAVID have been used to conduct the GO analysis of common TFs within three different categories like BP, MF and CC. Common statistically significant (p<0.05) ontological processes have been identified through DAVID.

Important biological processes like cellular response to interleukin-1, interleukin-6, Interferon-gamma-mediated signaling pathway, macrophage differentiation, negative regulation by host of viral transcription, cell proliferation, fat cell differentiation, interleukin-2 biosynthetic process, transcription from RNA polymerase II promoter, Positive regulation by host of viral transcription, interleukin-12 biosynthetic process, miRNA metabolic process, pri-miRNA transcription from RNA polymerase II promoter, transcription, Regulation of inflammatory response, miRNA mediated inhibition of translation are involved in the metabolic regulatory process of TFs.

Most of the TFs play significant roles in different MF like RNA polymerase II core promoter proximal region and distal enhancer sequence-specific DNA binding, core promoter binding, transcriptional repressor activity, chromatin DNA binding, transcriptional activator activity, RNA polymerase II transcription regulatory region sequence-specific binding, chromatin binding, Histone deacetylase activity, and ligand-activated sequence-specific DNA binding.

Various transcription factors localization in multiple CC like cytoplasm, nucleus, nuclear euchromatin, nuclear chromatin, Transcription factor complex, Sin3 complex, nucleoplasm, NuRD complex, ESC/E(Z) complex, and cytosol **(figure 5 and Supplementary Table 4A-4C)**.

**Figure 3.**
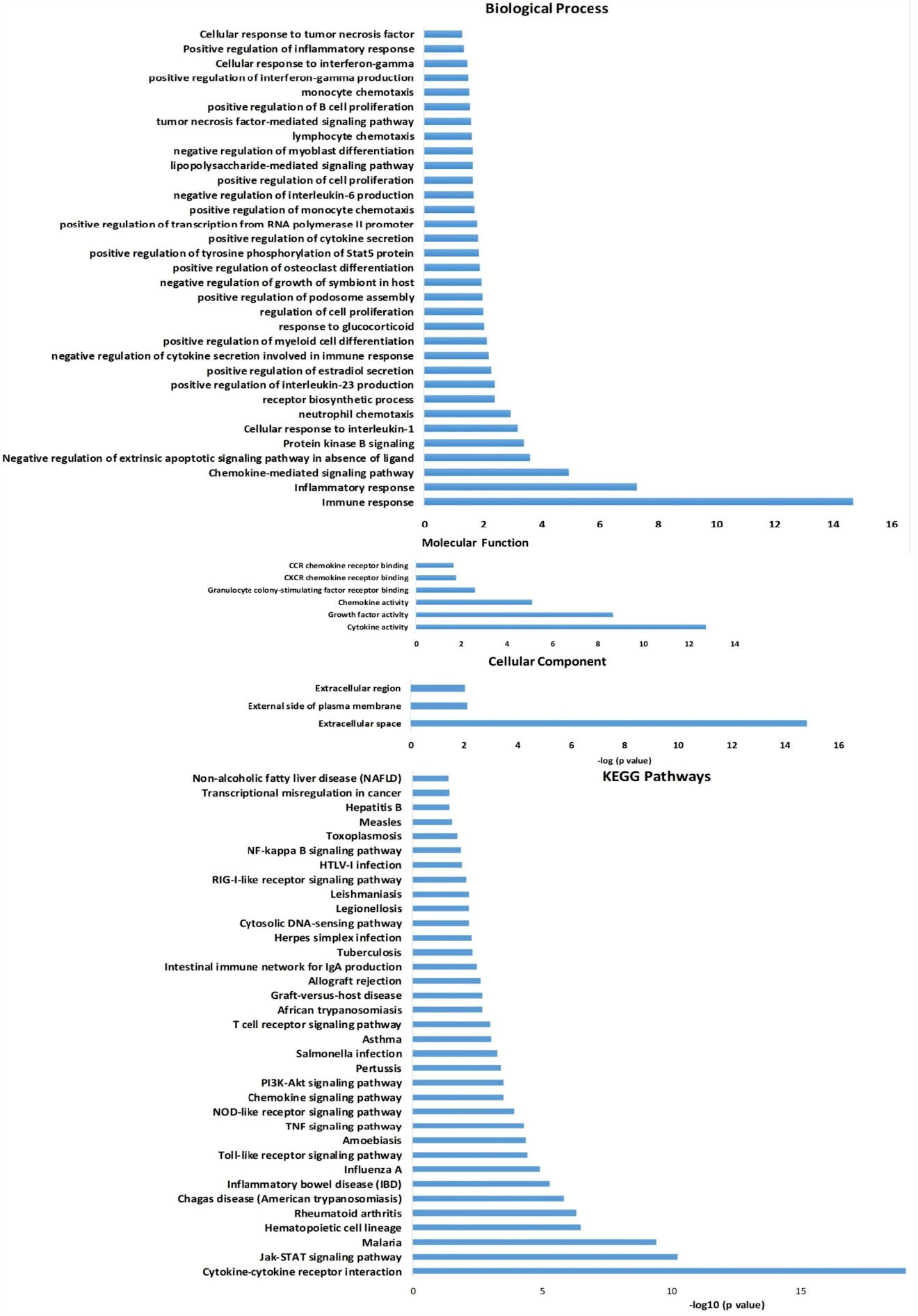
Enriched Gene Ontology terms of Cytokine storm genes obtained; Biological Process (BP), Molecular Function (MF), Cellular components (CC), KEGG Pathway.

**Figure 4.**
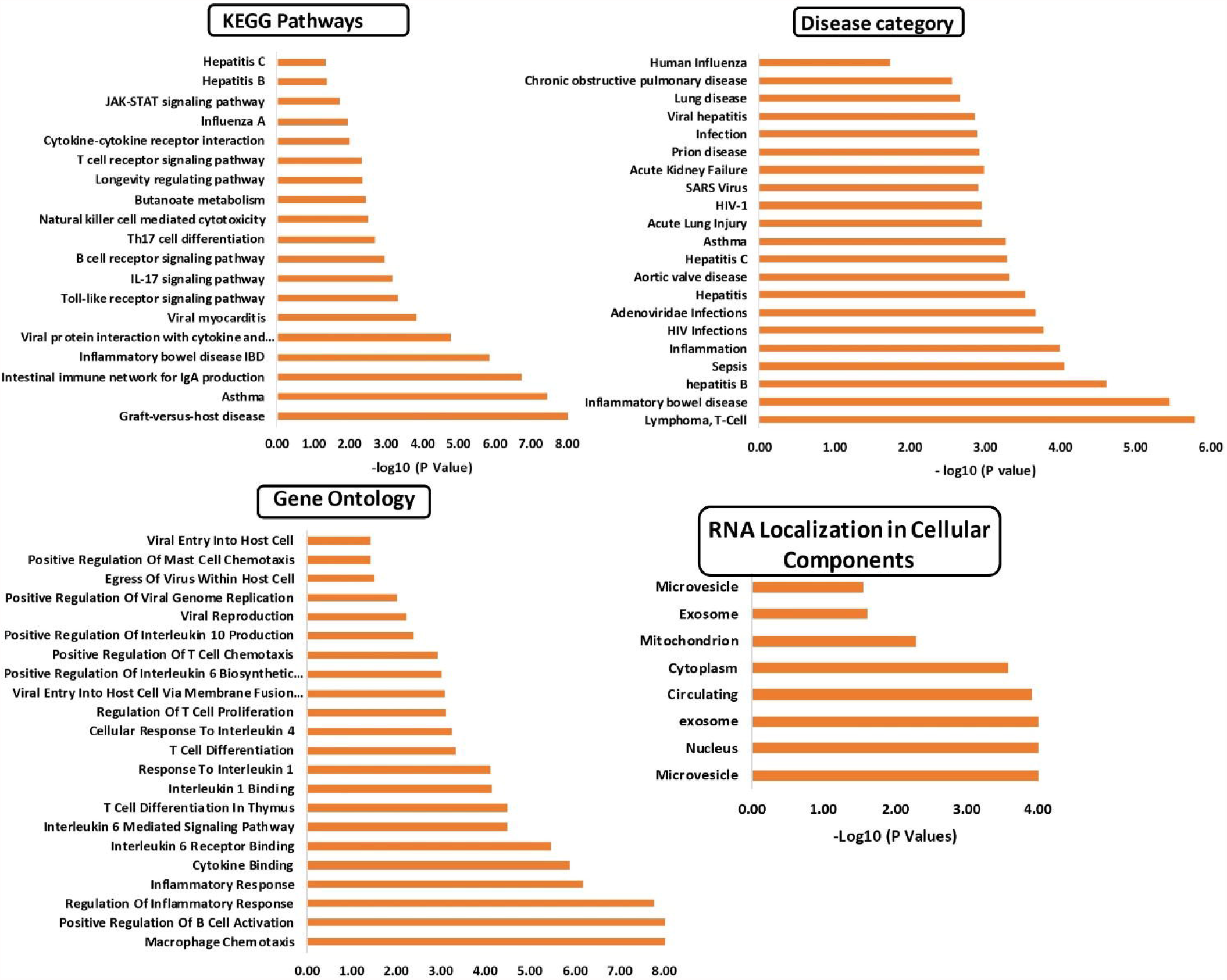
Common targeting MicroRNAs responsible for KEGG Pathway; RNA localization in cellular components (BP), Gene ontology (GO), Disease Category (DC).

**Figure 5.**
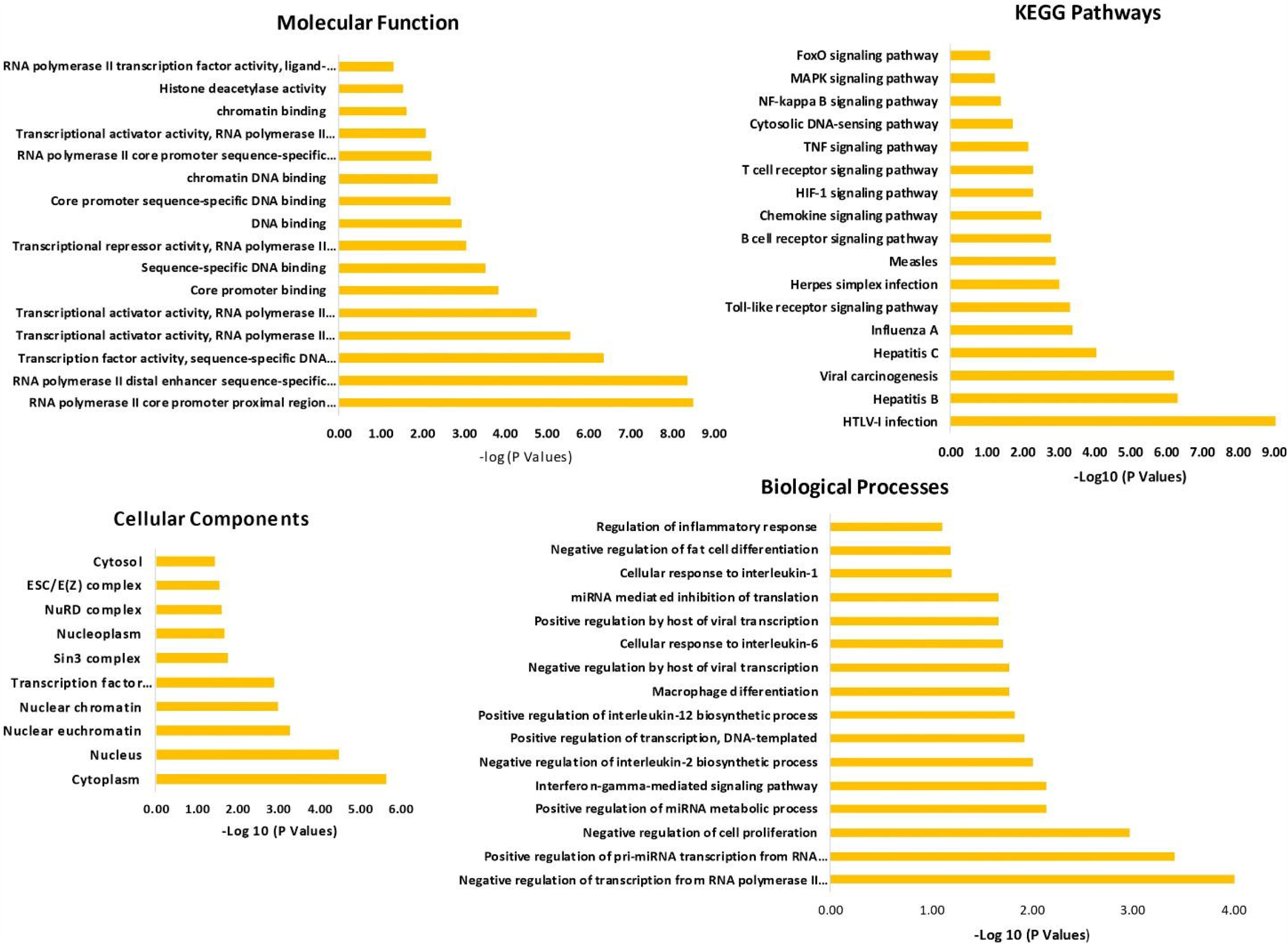
Enriched Gene Ontology terms of Cytokine storm genes obtained; KEGG Pathway; RNA localization in cellular components (BP), Gene ontology (GO), Disease Category (DC).

### 3.7 Common Pathway enrichment analysis of transcription factors

Kyoto Encyclopedia of Genes and Genomes (KEGG) pathway analysis of common transcription regulating genes identified above was carried out. A p-value<0.05 was set as the threshold for the significantly enriched pathways.

Crucial KEGG pathways involved in the common TFs were HTLV-I infection, Hepatitis B, Hepatitis C, Influenza A, Viral carcinogenesis, Herpes simplex infection, Measles, Toll-like receptor signaling pathway, B cell receptor signaling pathway, chemokine signaling pathway, HIF-1 signaling pathway, T cell receptor signaling pathway, TNF signaling pathway, cytosolic DNA-sensing pathway, NF-kappa B signaling pathway, MAPK signaling pathway, and FoxO signaling pathway. **(Supplementary Table 4D)**

### 3.8 Identification of common targeting MicroRNAs of CRIEGs and TFs

We identified the common MicroRNAs targeting CRIEGs and TFs from various microRNA databases like miRNet, TargetScan, miRDB, miRanda, miRWalk. During this data identification process, common CRIEGs and TFs multi-targeting 10 miRNAs-hsa-miR-106a-5p, hsa-miR-155-5p, hsa-miR-98-5p, hsa-miR-24-3p, hsa-miR-204-5p, hsa-miR-124-3p, hsa-miR-203a-3p, hsa-miR-335-5p, hsa-let-7c-5p, and hsa-miR-1-3p were obtained. All these miRNAs target the CRIEGs and its same targeting TFs **(Tables 1 & 2)**.

### 3.9 Disease category, RNA localization and Ontological analysis of frequent targeting MicroRNAs

We identified the microRNA enrichment analysis from two different databases MIENTURNET and miEA. We found out the localization of cellular components, miRNA-disease relationship and ontological functions of these important microRNAs. These ten MicroRNAs are found in different parts of the cell, such as microvesicle, nucleus, exosome, cytoplasm, and mitochondrion. These miRNAs also correlate in many diseases such as SARS, lymphoma, inflammatory bowel disease, hepatitis B, hepatitis C, asthma, Acute Lung Injury, sepsis, HIV, Adenoviridae infections, aortic valve disease, Acute Kidney Failure, prion disease, chronic obstructive pulmonary disease, and human influenza.

Involvement of miRNA in various GO terms such as macrophage chemotaxis, positive regulation of B-cell activation, regulation of inflammatory response cytokine binding, IL-6 receptor binding, IL-6 Mediated Signaling Pathway, T-cell differentiation in thymus, IL-1 binding, response To IL-1, cellular response to IL-4, regulation of T-cell proliferation, viral entry into host cell via membrane fusion with the plasma membrane, positive regulation of IL-6 biosynthetic process, T-cell chemotaxis, IL-10 production, mast cell chemotaxis viral reproduction, positive regulation of viral genome replication, viral entry into host cell. **(Figure 4 & Tables 5B-5D)**

### 3.10 Common Pathway enrichment analysis of MicroRNA

These ten crucial MicroRNAs regulate Graft-versus-host disease, asthma, intestinal immune network for IgA production, IBD, viral protein interaction with cytokine and cytokine receptor, viral myocarditis, Toll-like receptor signaling pathway, B-cell receptor signaling pathway, Th17 cell differentiation and signaling pathway, natural killer cell mediated cytotoxicity, Butanoate metabolism, longevity regulating pathway, T-cell receptor signaling pathway, cytokine-cytokine receptor interaction, Influenza A, JAK-STAT signaling pathway, Hepatitis B, Hepatitis C-that are responsible for progression of many metabolic and pathological regulatory process. **(Table 5A)**

### 3.11 PPIs network of CRIEGs, TFs and common targeting MicroRNAs interactions network with CRIEGs, TFs

With the help of Cytoscape, we created a network of common targeting miRNAs of all the 14 CRIEGs and their 32 TFs. These all play important correlations that affect the entire network due to the influence of an external specific virus which contributes to increase the severity of the disease. **(Table 1 & Figure 7, 8 & 9)**

**Figure 6.**
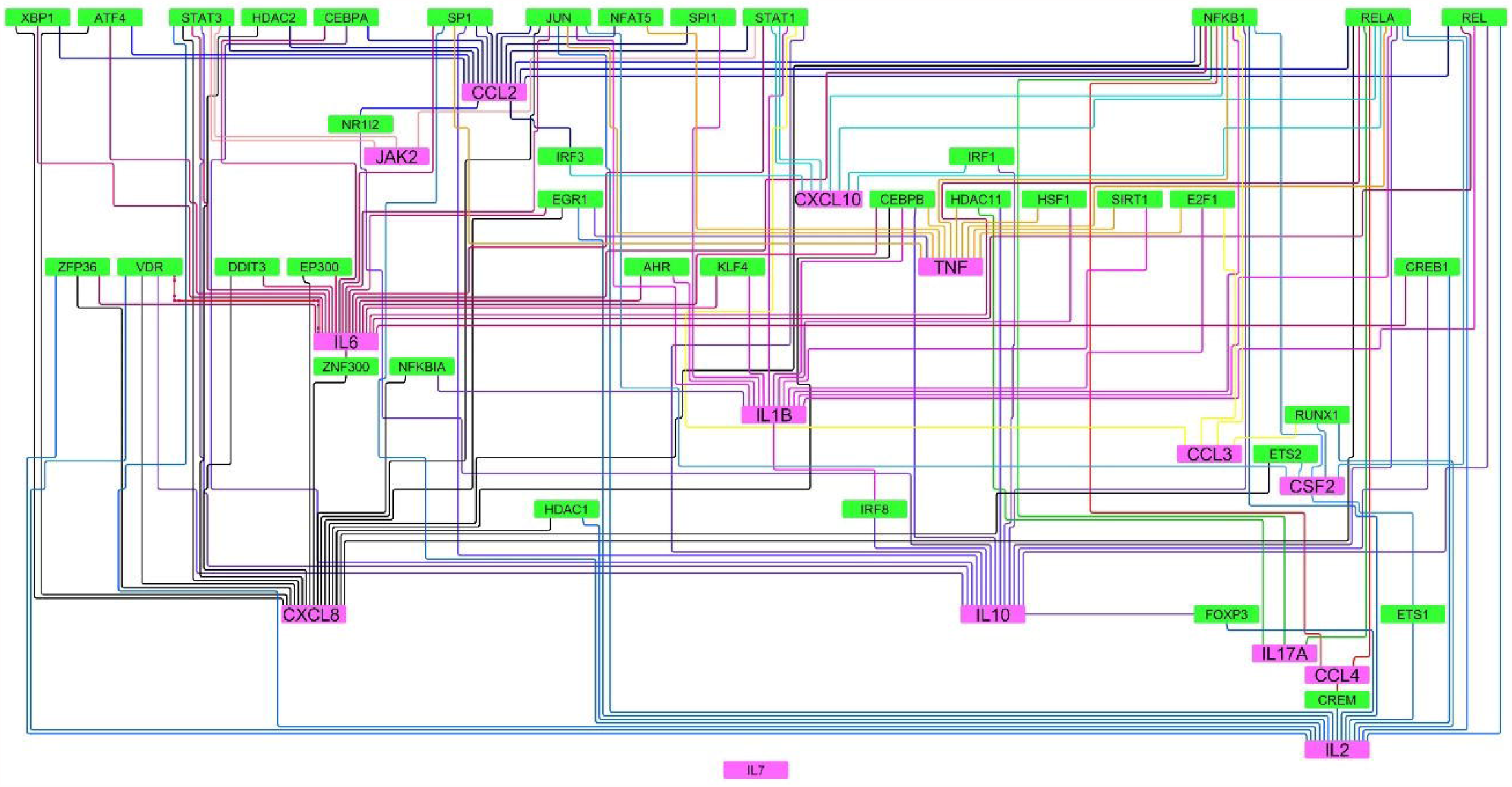
Show the PPI network of Cytokine and Transcription interaction network.

**Figure 7.**
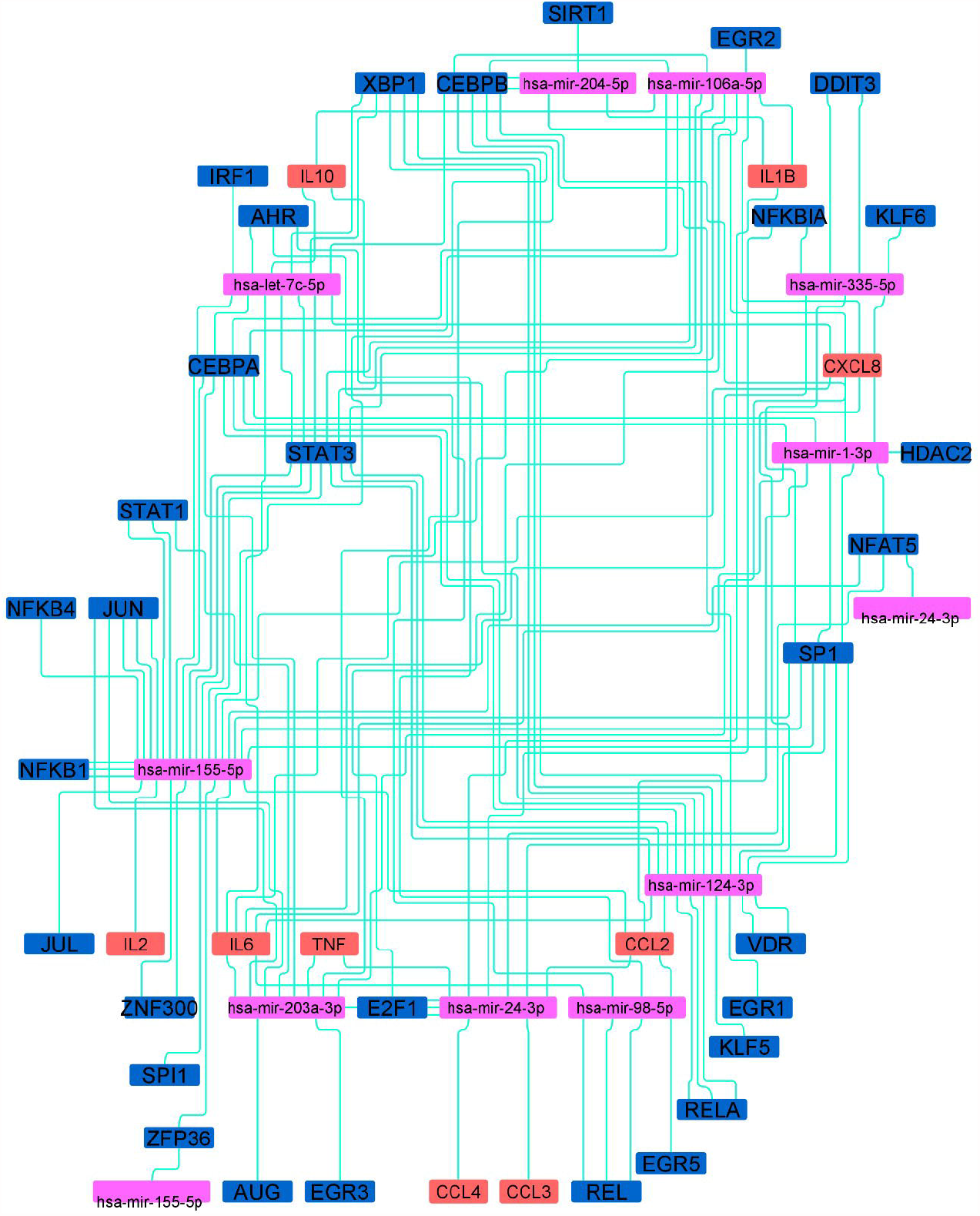
Show the Cytokine-Transcription factor and MicroRNAs interaction network.

**Figure 8.**
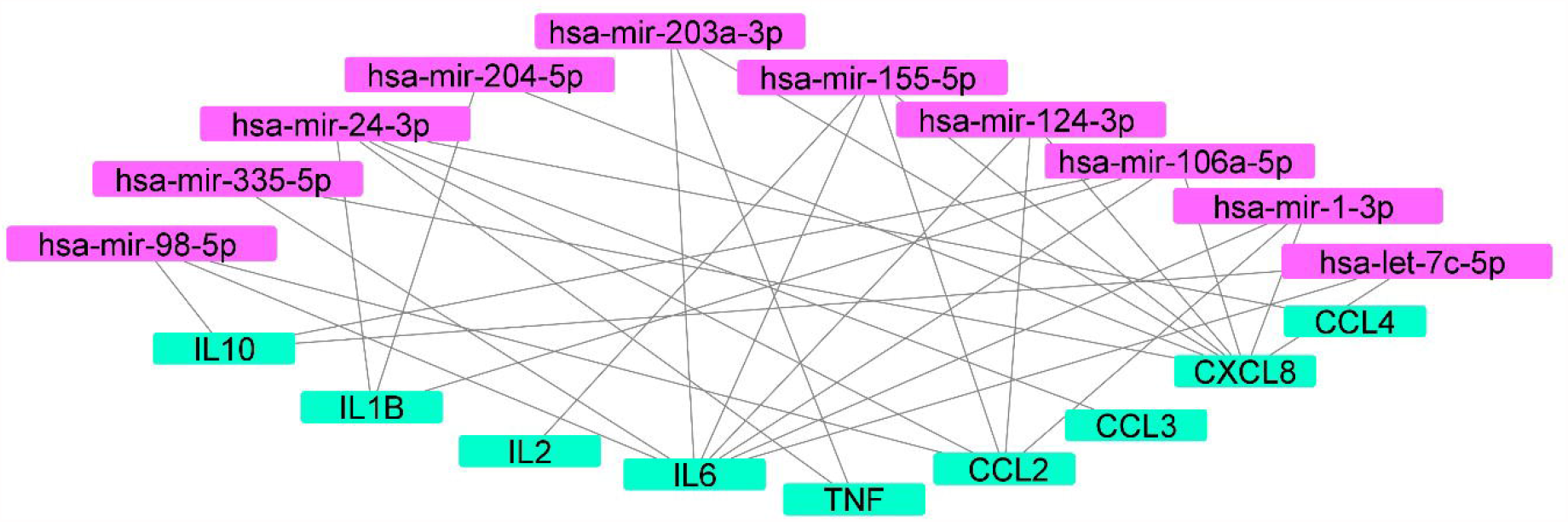
The Cytokine and MicroRNAs interaction network.

## 4. Discussion

MicroRNAs post-transcriptionally regulate the expression of target mRNA. RNA viruses are known to utilize the host miRNA machinery for their benefit. Hence, various studies have identified miRNAs as key players in the pathogenesis and therapeutics of viral diseases. Also, miRNAs can target viral genes as well as the host inflammatory machinery, as part of the host-pathogen interactions, to counter-act the impairing effects of infection (Ghosh *et al*. 2008). Demirci et al. have identified 67 different human miRNA that targets the spike protein of the SARS-CoV-2 virus (Saçar Demirci and Adan 2020). The inflammatory cascades involved in the pathophysiological pathways of COVID-19 are crucial in the development of complications in COVID-19. These pathogenetic pathways constitute, but are not limited to, receptor tyrosine kinases, the JAK/STAT pathway, TNF-α receptor and toll-like receptors, IL-6 and IFN-γ, cytokine storm, and macrophage activation (Yarmohammadi *et al*. 2020). Hence, exploring the regulatory networks of these inflammatory markers have the two-fold advantages of discovering the interconnected nature of these dysregulated pathways and unlocking the potential of novel mechanistic-based treatment strategies. However, a clear understanding of the miRNA response in SARS-CoV-2 is still elusive. Here, we have identified the miRNAs and transcription factors of the target mRNAs which provide the necessary insight into the genetic regulation of the inflammatory response in COVID-19.

Our in-silico analysis revealed ten miRNAs involved in the regulation of the common inflammatory genes and their transcription factors.

The miR-155-5p has been widely studied in viral inflammatory pathways. It is a regulator of the HCV-induced TLR3/NF-κB pathway mediated inflammatory response. Further, elevated circulating levels of miR-155 were also observed in HBV infection (Bala *et al*. 2012, Wang *et al*. 2015). The role of miR-155-5p in the cytokine response through the TLR4/NF-κB/miR-155-5p/SOCS-1 axis in monocyte-derived macrophages has been demonstrated in dengue (Arboleda *et al*. 2019). It has further been observed to be upregulated in B cells in EBV infection and in PBMC of HIV-1 infected patients (Gao *et al*. 2015, Dey *et al*. 2016). In JHMV-infected (a coronavirus) mice models, miR-155 enhanced the T-cell trafficking, cytokine secretion, and cellular effectors functions (Dickey *et al*. 2016). Woods et al. studied 1908 mature murine miRNA expressions in influenza A virus (IAV)-infected type II alveolar cells and miR-155-5p was showed to have the highest expression (Woods *et al*. 2020). In FeAE cells, miR-155-5p expression induced IL-6 and IP-10 production, which is responsible for recruitment of leukocytes (McAdams *et al*. 2015). It also regulates the NF-κB and MAPK signaling pathways (Shi *et al*. 2020). In our analysis, we found miR-155-5p to be one of the ten identified miRNAs that possibly regulates the cytokine expression and triggers inflammatory response in COVID-19. Further, miR-155-5p was found to affect the expression of multiple transcription factors including CEBPA, JUN, NFAT5, NFKB1, SP1, SPI1, STAT1, STAT3, CEBPB, ZP36, ZNF300, ZFP36.

Another targeting miRNA identified in this study, miR-124-3p, was observed to be downregulated in JEV-infected human neural stem/progenitor cells (Mukherjee *et al*. 2019). A mice model showed downregulation in miR-124-3p expression in ARDS. Treatment with miR-124-3p agomir attenuated the pulmonary injury and the levels of pro-inflammatory cytokines IL-6 and TNF-α by directly targeting p65, thus showing promise in in-vitro management of pulmonary injury (Liang *et al*. 2020, p. 65).

Yet another miR-203a has been demonstrated to have an antagonistic role in foot-and-mouth disease virus (FMDV) infection (Gutkoska *et al*. 2017). This miRNA was further studied in IAV infection where an upregulated miR-203a modulated the antiviral response by targeting DR1 gene (Zhang *et al*. 2018, p. 1). However, further studies are needed to consolidate the its role in corona-virus infections. Our in-silico analysis showed miR-203a targets transcription factors NFkB1, RELA, CREBPB, ATF4, ETS1 and 2.

miR-335-5p had the most predicted targets in the response against the Porcine Reproductive and Respiratory Syndrome Virus (PRRSV) of alveolar macrophages. Contrastingly, no effects on the cytokine expression was observed in this study (Dhorne-Pollet *et al*. 2019). This may be attributed to the low level of expression of miR-335-5p in most tissues, which renders its effect to be negligible despite the abundant number of predicted targets (The FANTOM Consortium *et al*. 2017). However, miR-24-3p facilitated PRRSV replication via suppression of heme oxygenase-1 (HO-1) (Xiao *et al*. 2015), and HO-1 has been reported to play role in anti-viral activity in several viral infections including HIV, hepatitis C virus, hepatitis B virus, enterovirus 71, influenza virus, respiratory syncytial virus, dengue virus, and Ebola virus (Espinoza *et al*. 2017, p. 1). Thus a high expression of miR-24-3p may be pathognomic for the worsening of viral infection.

Another targeting miRNA identified in this study, hsa-let-7c-5p, directly affects ACE2 and TMPRSS2; two key players in the SARS-CoV-2 infection (Chauhan *et al*. 2020). In rhabdomyosarcoma cells, hsa-let-7c-5p promoted the replication of enterovirus 71 (EV71) by inhibition of the MAPK4K expression (Zhou *et al*. 2017). Overexpression of miR-let-7c attenuated the replication of HCV through HO-1 induction (Chen *et al*. 2019). A differential expression of hsa-let-7 in-silico as a CRIEG indicates it has a role in immunomodulation in COVID-19.

Collectively, these studies demonstrate that mainly ten microRNAs (hsa-miR-106a-5p, hsa-miR-155-5p, hsa-miR-98-5p, hsa-miR-24-3p, hsa-miR-204-5p, hsa-miR-124-3p, hsa-miR-203a-3p, hsa-miR-335-5p, hsa-let-7c-5p, hsa-miR-1-3p) regulate the role of inflammatory mechanism in viral infection. Our in-silico analysis points towards a similar potential regulatory role of miRNA in SARS-CoV-2 mediated inflammatory cascades. Many of the target miRNA found in this study, namely miR-106a-5p, miR-1-3p, miR-98-5p, miR-24-3p, and miR-204-5p have been observed to orchestrate the gene expression of IL-1β, IL-6, IL-10, IFNγ, IL-2, and IL-17 through TFs such as ERK, STAT1, and STAT3 (Ye *et al*. 2014, Srivastava *et al*. 2017, Shen *et al*. 2019, Xiu *et al*. 2020).

Although most of these data are derived from cancer and transplantation studies, a similar mechanism of regulation of expression may be functional in the case of COVID-19. The disease severity in COVID-19 had been associated with an influx of innate immune cells and inflammatory cytokines (Hadjadj *et al*. 2020, Yale IMPACT Team *et al*. 2020, p. 19). The cytokine storm in COVID-19 leads to lung injury, multiple organ failure and had poor prognosis (Jose and Manuel 2020, Mehta *et al*. 2020). TNF-α and IFN-γ together had showed to incite the cells to PANoptosis, inflammatory cell death involving the components of pyroptosis, apoptosis, and necroptosis. Further, JAK/STAT1/IRF1 axis was also involved in regulation of inflammatory cell death due to PANoptosis (Karki *et al*. 2020). Further, IL-6 have also been shown to activate the Janus kinase-Signal Transducer and Activator of Transcription (JAK-STAT) pathway leading to immune activation (Luo *et al*. 2020, p. 19).

Apart from the above mentioned transcription factors, NF-κB also plays a crucial role in poor prognosis of severe COVID-19 disease. NF-κB leads to accelerated inflammatory response with increased secretion of TNF-α and IL-6. This auto-amplified pro-inflammatory loop with impaired type I IFN response culminates in Viral replication within the lungs and tissue damage (Hadjadj *et al*. 2020). Recent studies have demonstrated the up regulation of various miRNAs in COVID-19 patients thus confirming our prediction. In comparison to healthy controls, miRNAs, including miR-21, miR-155, miR-208a and miR-499, had been demonstrated to be up-regulated in the COVID-19 patients (Mahesh and Biswas 2019, Garg *et al*. 2021). The varying functional significance and organ specificty of these upregulated miRNAs,i.e, miR-155 (inflammatory miRNA), miR-208a (heart-muscle specific) and miR-499 (muscle function) and miR-21 (fibrosis-associated) denotes the involvement of multiple pathways and organs in the pathophysiology of COVID-19 (van Rooij *et al*. 2007, 2009, Thum *et al*. 2008). MicroRNAs such as miR-125b, miR-138, miR-199a and miR-21 are also responsible for cytokine storms in the acute respiratory distress syndrome and COPD(Guterres *et al*. 2020). In addition, miR-26a-5p, miR-29b-3p, and miR-34a-5p have been shown to be involved in endothelial dysfunction and inflammatory response in patients with SARS–CoV-2 infection(Centa *et al*. 2021). Further, the upregulation miRNAs in post-COVID-19 complications such as (miR-21, miR-155, miR-208a and miR-499) in chronic myocardial damage and inflammation(Garg *et al*. 2021), (let-7b-3p, miR-29a-3p, miR-146a-3p and miR-155-5p) in post-acute COVID-19 phase; also indicates the extent of the alteration of these pathways in Post-COVID-19 sequelae. The downregulation of these miRNAs may be targeted to improve acute symptoms and distress by regulating the production of pro-inflammatory cytokines and apoptotic proteins (Guterres *et al*. 2020). Apart from the effect on host cellular pathways, miRNAs can also inhibit the viral infectivity by different ways like blocking the viral replication, cellular receptors and the function of viral proteins in SARS-CoV-2 (Fani *et al*. 2021). MicroRNAs such as miR-21-3p, miR-195-5p, miR-16-5p, miR-3065-5p, miR-424-5p and miR-421 potentially regulate the infectivity of viruses belonging to human coronavirus family through direct binding to the viral genome (Chan *et al*. 2020). Interestingly, newer miRNA-based therapy including antimiR-18 and antimir-125b, which potentially targets ACE2-related genes, have been proposed for nephropathy associated with COVID-19 (Widiasta *et al*. 2020).

We have identified other target CRIEGs of transcription factors STAT1, IRF1 & NF-κB and the miRNAs regulating their expression. This would help to better understand the cross talk and regulation of various cytokines in COVID-19 and the possible role played by them in regulation of inflammatory cell death leading to multiple organ failure. The main limitation of this study is that the data is derived from the publicly available databases, and needs experimental substantiation to prove its clinical efficacy. Moreover, our study highlights the interaction and the pathways concerning the miRNA, immune-expressed and TFs. Such expression data of all these three entities together are not available in COVID-19 patients or in-vitro models, which can establish a better understanding of the mechanisms involved.

## 5. Conclusion

The present study identifies an in-silico representation of a network involving miRNAs (hsa-miR-106a-5p, hsa-miR-155-5p, hsa-miR-98-5p, hsa-miR-24-3p, hsa-miR-204-5p, hsa-miR-124-3p, hsa-miR-203a-3p, hsa-miR-335-5p, hsa-let-7c-5p, hsa-miR-1-3p), CRIEGs (CCL2, CCL4, CXCL10, CXCL8, IL6, IL7, JAK2, TNF), and TF (AHR, CREM, DDIT3, E2F1, EGR1, EP300, ESR1, ETS2, HDAC1, HDAC2, IRF1, JUN, KLF4, NFAT5, NFKB1, NFKBIA, REL, RUNX1, SIRT1, SP100, SP140L, STAT1, XBP1, ZFP36) which take part in the inflammatory response in COVID-19. This study has identified the CRIEGs and miRNA, the interactions between them, which are potentially critical and can be studied further for the development of targeted therapeutic strategies. The data can also be used in exploring novel pathways, which occur following SARS-CoV-2 infection. However, the data needs to be experimentally validated in vitro and in vivo.

## Data Availability

The authors confirm that the data supporting the findings of this study are available within the manuscript and its supplementary materials.

## 7. Acknowledgements

The authors thanks to Dr. Dipayan Roy (MD Biochemistry) for language editing and review of the manuscript.

