## Supplemental File for "Micro RNA-based regulation of genomics and transcriptomics of inflammatory cytokines in COVID-19"

**Supplimentry File:**

**Table: 1.** Inflammatory cytokine responsible for cytokine storm in Covid-19

| Sr. No. | Gene Symbol | Symbol | Gene | References |
| --- | --- | --- | --- | --- |
| 1 | IL-1b | IL1B | Interleukin-1 beta; | (1) (2) (3) |
| 2 | IL-2 | IL2 | Interleukin-2; | (4) (2) (5) (6) (7) (8) |
| 3 | IL-7 | IL7 | Interleukin-7; | (1) |
| 4 | IL-8 | CXCL8 | Interleukin-8; | (1,9) (2) |
| 5 | IL-9 | IL9 | Interleukin-9; |  |
| 6 | IL-10 | IL10 | Interleukin-10; | (1) (10) (9,11–13) (4) (2) (5) (14) (6) (7) (8) |
| 7 | IL-17 | IL17A | Interleukin-17A; | (1) (7,10) (3) |
| 8 | G-CSF | CSF3 | Granulocyte colony-stimulating factor; |  |
| 9 | GM-CSF | CSF2 | Granulocyte-macrophage colony-stimulating factor; |  |
| 10 | IFN-gamma | JAK2 | Tyrosine-protein kinase JAK2; | (1) (10) (12) (5,14) (6) (7) |
| 11 | TNF-alpha | TNF | Tumor necrosis factor; | (1) (10) (9,11) (4) (2) (5) (14) (6) (7) (3) |
| 12 | CXCL10 | CXCL10 | C-X-C motif chemokine 10; |  |
| 13 | MCP1 | CCL2 | C-C motif chemokine 2; | (15,16) |
| 14 | MIP1A | CCL3 | C-C motif chemokine 3; | (15,16) |
| 15 | MIP1B | CCL4 | C-C motif chemokine 4; | (15,16) |
| 16 | IL-6 | IL6 | Interleukin-6; | (1) (3) (2) (17) (10) (9,11,12) (18) (19)(7,20) (4) (5,21) (22) (14) (6) (8,13) |

Table 2: Common (String and DAVID) Gene Ontology and Functional Enrichment.

| Molecular Function | Cellular Component | Common Biological Process | KEGG Pathway |
| --- | --- | --- | --- |
| CCR chemokine receptor binding | extracellular space | humoral immune response | Herpes simplex infection |
| Chemokine activity | external side of plasma membrane | monocyte chemotaxis | NOD-like receptor signaling pathway |
| Growth factor activity |  | negative regulation of myoblast differentiation | Malaria |
| CXCR chemokine receptor binding |  | positive regulation of B cell proliferation | Tuberculosis |
| Cytokine activity |  | receptor biosynthetic process | African trypanosomiasis |
|  |  | negative regulation of cytokine secretion involved in immune response | NF-kappa B signaling pathway |
|  |  | positive regulation of osteoclast differentiation | Influenza A |
|  |  | positive regulation of cytokine secretion | Leishmaniasis |
|  |  | cellular response to interferon-gamma | Asthma |
|  |  | response to glucocorticoid | HTLV-I infection |
|  |  | negative regulation of growth of symbiont in host | Cytosolic DNA-sensing pathway |
|  |  | lipopolysaccharide-mediated signaling pathway | Non-alcoholic fatty liver disease (NAFLD) |
|  |  | chemokine-mediated signaling pathway | TNF signaling pathway |
|  |  | response to lipopolysaccharide | Intestinal immune network for IgA production |
|  |  | positive regulation of inflammatory response | Chemokine signaling pathway |
|  |  | positive regulation of interleukin-23 production | Hepatitis B |
|  |  | immune response | Toxoplasmosis |
|  |  | positive regulation of GTPase activity | Amoebiasis |
|  |  | positive regulation of myeloid cell differentiation | Toll-like receptor signaling pathway |
|  |  | tumor necrosis factor-mediated signaling pathway | Legionellosis |
|  |  | protein kinase B signaling | RIG-I-like receptor signaling pathway |
|  |  | inflammatory response | Inflammatory bowel disease (IBD) |
|  |  | cellular response to lipopolysaccharide | Graft-versus-host disease |
|  |  | cellular response to interleukin-1 | Rheumatoid arthritis |
|  |  | negative regulation of interleukin-6 production | Chagas disease (American trypanosomiasis) |
|  |  | positive regulation of interferon-gamma production | Hematopoietic cell lineage |
|  |  | cellular response to tumor necrosis factor | Pertussis |
|  |  | positive regulation of podosome assembly | Transcriptional misregulation in cancer |
|  |  | negative regulation of extrinsic apoptotic signaling pathway in absence of ligand | Salmonella infection |
|  |  | adaptive immune response | Cytokine-cytokine receptor interaction |
|  |  | neutrophil chemotaxis | T cell receptor signaling pathway |
|  |  |  | Allograft rejection |
|  |  |  | PI3K-Akt signaling pathway |
|  |  |  | Autoimmune thyroid disease |
|  |  |  | Type I diabetes mellitus |
|  |  |  | Jak-STAT signaling pathway |

**Table 3A:** Molecular Function of cytokine genes.

| Term | PValue | Genes | Fold Enrichment | FDR | Negative LOG (Pvalues) |
| --- | --- | --- | --- | --- | --- |
| Cytokine activity | 0 | CSF3, CSF2, IL17A, IL6, TNF, IL7, IL10, IL2 | 60.19 | 0 | 12.79 |
| Growth factor activity | 0 | CSF3, CSF2, IL6, IL7, IL2 | 94.7 | 0 | 8.71 |
| Chemokine activity | 0 | CCL2, CXCL8, CCL4, CXCL10 | 92.84 | 0.01 | 5.13 |
| Granulocyte colony-stimulating factor receptor binding | 0.002 | CSF3 | 789.14 | 1.95 | 2.63 |
| CXCR chemokine receptor binding | 0.016 | CXCL8, CXCL10 | 112.73 | 12.89 | 1.79 |
| CCR chemokine receptor binding | 0.021 | CCL2, CCL4 | 87.68 | 16.26 | 1.68 |

**Table 3B:** KEGG Pathways of cytokine genes.

| Term | PValue | Genes | Fold Enrichment | FDR | Negative log 10 P Values |
| --- | --- | --- | --- | --- | --- |
| Cytokine-cytokine receptor interaction | 0 | CSF3, CSF2, IL17A, IL6, TNF, CCL2, IL7, IL9, CXCL8, CCL4, IL10, IL2, CXCL10 | 32.5 | 0 | 19.08 |
| Jak-STAT signaling pathway | 0 | CSF3, CSF2, IL6, IL7, IL9, JAK2, IL10, IL2 | 30 | 0 | 10.25 |
| Malaria | 0 | CSF3, IL6, CCL2, TNF, CXCL8, IL10 | 62.8 | 0 | 9.45 |
| Hematopoietic cell lineage | 0 | CSF3, CSF2, IL6, TNF, IL7 | 35 | 0 | 6.5 |
| Rheumatoid arthritis | 0 | CSF2, IL17A, IL6, CCL2, TNF, CXCL8 | 32.6 | 0 | 6.34 |
| Chagas disease (American trypanosomiasis) | 0 | IL6, CCL2, TNF, CXCL8, IL10, IL2 | 26.2 | 0 | 5.87 |
| Inflammatory bowel disease (IBD) | 0 | IL17A, IL6, TNF, IL10, IL2 | 38.3 | 0 | 5.31 |
| Influenza A | 0 | IL6, CCL2, TNF, CXCL8, JAK2, CXCL10 | 16.8 | 0 | 4.91 |
| Toll-like receptor signaling pathway | 0 | IL6, TNF, CXCL8, CCL4, CXCL10 | 23.1 | 0 | 4.44 |
| Amoebiasis | 0 | CSF2, IL6, TNF, CXCL8, IL10 | 22.4 | 0 | 4.39 |
| TNF signaling pathway | 0 | CSF2, IL6, CCL2, TNF, CXCL10 | 21.6 | 0 | 4.32 |
| NOD-like receptor signaling pathway | 0 | IL6, CCL2, TNF, CXCL8 | 38.1 | 0.1 | 3.95 |
| Chemokine signaling pathway | 0 | CCL2, CXCL8, JAK2, CCL4, CXCL10 | 13.5 | 0.3 | 3.53 |
| PI3K-Akt signaling pathway | 0 | CSF3, IL6, IL7, JAK2, IL2 | 8.6 | 0.3 | 3.52 |
| Pertussis | 0 | IL6, TNF, CXCL8, IL10 | 25.6 | 0.4 | 3.44 |
| Salmonella infection | 0 | CSF2, IL6, CXCL8, CCL4 | 23 | 0.5 | 3.31 |
| Asthma | 0 | TNF, IL9, IL10 | 60.9 | 0.9 | 3.04 |
| T cell receptor signaling pathway | 0 | CSF2, TNF, IL10, IL2 | 18.1 | 1 | 3 |
| African trypanosomiasis | 0 | IL6, TNF, IL10 | 41.2 | 2 | 2.7 |
| Graft-versus-host disease | 0 | IL6, TNF, IL2 | 41.2 | 2 | 2.7 |
| Allograft rejection | 0 | TNF, IL10, IL2 | 37.8 | 2.4 | 2.62 |
| Intestinal immune network for IgA production | 0 | IL6, IL10, IL2 | 32.6 | 3.2 | 2.49 |
| Tuberculosis | 0 | IL6, TNF, JAK2, IL10 | 10.7 | 4.5 | 2.35 |
| Herpes simplex infection | 0.01 | IL6, CCL2, TNF, JAK2 | 10.3 | 5.1 | 2.29 |
| Cytosolic DNA-sensing pathway | 0.01 | IL6, CCL4, CXCL10 | 23 | 6.3 | 2.2 |
| Legionellosis | 0.01 | IL6, TNF, CXCL8 | 22.6 | 6.5 | 2.18 |
| Leishmaniasis | 0.01 | TNF, JAK2, IL10 | 22.6 | 6.5 | 2.18 |
| RIG-I-like receptor signaling pathway | 0.01 | TNF, CXCL8, CXCL10 | 20.6 | 7.7 | 2.11 |
| HTLV-I infection | 0.01 | CSF2, IL6, TNF, IL2 | 7.5 | 11.5 | 1.92 |
| NF-kappa B signaling pathway | 0.01 | TNF, CXCL8, CCL4 | 15.7 | 12.6 | 1.88 |
| Toxoplasmosis | 0.02 | TNF, JAK2, IL10 | 13.3 | 16.9 | 1.74 |
| Measles | 0.03 | IL6, JAK2, IL2 | 10.6 | 24.9 | 1.56 |
| Hepatitis B | 0.04 | IL6, TNF, CXCL8 | 9.3 | 30.8 | 1.45 |
| Transcriptional misregulation in cancer | 0.04 | CSF2, IL6, CXCL8 | 9.2 | 31.1 | 1.44 |
| Non-alcoholic fatty liver disease (NAFLD) | 0.04 | IL6, TNF, CXCL8 | 8.8 | 33.7 | 1.4 |

**Table 3C:** Biological Process of cytokine genes.

| Term | PValue | Genes | Fold Enrichment | FDR | Negative log10 (P Value) |
| --- | --- | --- | --- | --- | --- |
| Immune response | 0 | CSF3, CSF2, IL6, IL7, IL9, CXCL8, CCL4, IL10, IL2, CXCL10 | 42.1 | 0 | 14.72 |
| Inflammatory response | 0 | IL17A, CCL2, CXCL8, JAK2, CCL4, IL10, CXCL10 | 28.37 | 0 | 7.31 |
| Chemokine-mediated signaling pathway | 0 | CCL2, CXCL8, CCL4, CXCL10 | 81.93 | 0.01 | 4.95 |
| Negative regulation of extrinsic apoptotic signaling pathway in absence of ligand | 0 | CSF2, TNF, IL7 | 119.66 | 0.3 | 3.62 |
| Protein kinase B signaling | 0 | CSF2, CCL2, TNF | 94.73 | 0.49 | 3.42 |
| Cellular response to interleukin-1 | 0 | IL17A, CCL2, CCL4 | 73.34 | 0.81 | 3.19 |
| neutrophil chemotaxis | 0 | CCL2, CXCL8, CCL4 | 56.84 | 1.35 | 2.97 |
| receptor biosynthetic process | 0 | TNF, IL10 | 505.24 | 4.6 | 2.43 |
| positive regulation of interleukin-23 production | 0 | CSF2, IL17A | 505.24 | 4.6 | 2.43 |
| positive regulation of estradiol secretion | 0 | IL6, TNF | 378.93 | 6.09 | 2.31 |
| negative regulation of cytokine secretion involved in immune response | 0.01 | TNF, IL10 | 303.15 | 7.55 | 2.21 |
| positive regulation of myeloid cell differentiation | 0.01 | CSF3 | 252.62 | 8.99 | 2.13 |
| response to glucocorticoid | 0.01 | TNF, IL10 | 216.53 | 10.41 | 2.07 |
| regulation of cell proliferation | 0.01 | CXCL8, JAK2, CXCL10 | 19.11 | 11 | 2.04 |
| positive regulation of podosome assembly | 0.01 | CSF2, TNF | 189.47 | 11.81 | 2.01 |
| negative regulation of growth of symbiont in host | 0.01 | TNF, IL10 | 168.41 | 13.18 | 1.96 |
| positive regulation of osteoclast differentiation | 0.01 | IL17A, TNF | 151.57 | 14.53 | 1.91 |
| positive regulation of tyrosine phosphorylation of Stat5 protein | 0.01 | CSF2, IL2 | 137.79 | 15.87 | 1.87 |
| positive regulation of cytokine secretion | 0.01 | TNF, IL10 | 126.31 | 17.18 | 1.83 |
| positive regulation of transcription from RNA polymerase II promoter | 0.01 | IL17A, TNF, IL10, IL2 | 6.95 | 17.38 | 1.83 |
| positive regulation of monocyte chemotaxis | 0.02 | CCL2, CXCL10 | 101.05 | 20.99 | 1.74 |
| negative regulation of interleukin-6 production | 0.02 | TNF, IL10 | 89.16 | 23.44 | 1.68 |
| positive regulation of cell proliferation | 0.02 | CSF3, CXCL10 | 12.16 | 24.19 | 1.67 |
| lipopolysaccharide-mediated signaling pathway | 0.02 | CCL2, TNF | 84.21 | 24.63 | 1.66 |
| negative regulation of myoblast differentiation | 0.02 | TNF, CXCL10 | 84.21 | 24.63 | 1.66 |
| lymphocyte chemotaxis | 0.02 | CCL2, CCL4 | 79.78 | 25.81 | 1.64 |
| tumor necrosis factor-mediated signaling pathway | 0.03 | TNF, JAK2 | 72.18 | 28.1 | 1.59 |
| positive regulation of B cell proliferation | 0.03 | IL7, IL2 | 68.9 | 29.23 | 1.57 |
| monocyte chemotaxis | 0.03 | CCL2, CCL4 | 63.16 | 31.42 | 1.54 |
| positive regulation of interferon-gamma production | 0.03 | TNF, IL2 | 60.63 | 32.49 | 1.52 |
| Cellular response to interferon-gamma | 0.03 | CCL2, CCL4 | 54.13 | 35.6 | 1.47 |
| Positive regulation of inflammatory response | 0.04 | CCL2, CCL4 | 43.31 | 42.32 | 1.37 |
| Cellular response to tumor necrosis factor | 0.05 | CCL2, CCL4 | 36.97 | 47.52 | 1.31 |

**Table 3D:** Cellular Components of cytokine genes

| Term | PValue | Genes | Fold Enrichment | FDR | Negative log p value |
| --- | --- | --- | --- | --- | --- |
| Extracellular space | 0 | CSF3, CSF2, IL17A, IL6, TNF, CCL2, IL7, IL9, CXCL8, CCL4, IL10, IL2, CXCL10 | 15.71846 | 9.10E-13 | 14.86493 |
| External side of plasma membrane | 0.0073 | IL17A, TNF, CXCL10 | 21.38833 | 4.876031 | 2.134776 |
| Extracellular region | 0.0089 | IL6, IL7, CCL4, CXCL10 | 8.40819 | 5.880678 | 2.05147 |

**Table: 4A** Cellular components of Transcription Factor:

| Term | Count | PValue | Genes | Fold Enrichment | FDR |
| --- | --- | --- | --- | --- | --- |
| Cytoplasm | 15 | 0 | ZFP36, EGR1, CREM, ESR1, FOXP3, STAT1, STAT3, EP300, HDAC2, REL, HSF1, ETS2, IRF1, RUNX1, KLF4 | 3.874215 | 0.002002 |
| Nucleus | 13 | 0 | ZFP36, EGR1, CREM, RELA, ESR1, NFKBIA, FOXP3, STAT1, AHR, STAT3, NR1I2, REL, ZNF300 | 3.70135 | 0.030346 |
| Nuclear euchromatin | 3 | 0 | JUN, SIRT1, KLF4 | 84.37179 | 0.469105 |
| Nuclear chromatin | 4 | 0 | SPI1, IRF1, ESR1, STAT3 | 19.28498 | 0.898893 |
| Transcription factor complex | 4 | 0 | EP300, CREM, JUN, RELA | 17.91967 | 1.110473 |
| Sin3 complex | 2 | 0.02 | HDAC2, HDAC1 | 112.4957 | 14.33145 |
| Nucleoplasm | 7 | 0.02 | HSF1, JUN, ETS2, NFKB1, RUNX1, STAT3, KLF4 | 2.985354 | 17.54299 |
| NuRD complex | 2 | 0.02 | HDAC2, HDAC1 | 77.88166 | 20.02587 |
| ESC/E(Z) complex | 2 | 0.03 | HDAC2, SIRT1 | 67.49744 | 22.73014 |
| Cytosol | 5 | 0.04 | ZFP36, HDAC1, RELA, NFKBIA, NFKB1 | 3.777842 | 28.15102 |

**Table: 4B** Biological Process of Transcription Factor:

| Term | PValue | Genes | Fold Enrichment | FDR |
| --- | --- | --- | --- | --- |
| Negative regulation of transcription from RNA polymerase II promoter | 0 | ZFP36, HDAC2, EP300, ESR1, FOXP3, SIRT1, DDIT3 | 8.75 | 0.13 |
| Positive regulation of pri-miRNA transcription from RNA polymerase II promoter | 0 | JUN, SPI1, STAT3 | 98.69 | 0.52 |
| Negative regulation of cell proliferation | 0 | HSF1, JUN, IRF1, STAT3, KLF4 | 10.39 | 1.49 |
| Positive regulation of miRNA metabolic process | 0.01 | RELA, NFKB1 | 263.19 | 9.69 |
| Interferon-gamma-mediated signaling pathway | 0.01 | IRF1, STAT1 | 263.19 | 9.69 |
| Negative regulation of interleukin-2 biosynthetic process | 0.01 | ZFP36, FOXP3 | 197.39 | 12.71 |
| Positive regulation of transcription, DNA-templated | 0.01 | REL, ETS2, ESR1, STAT1 | 8.06 | 15.06 |
| Positive regulation of interleukin-12 biosynthetic process | 0.01 | RELA, IRF1 | 131.59 | 18.45 |
| Macrophage differentiation | 0.02 | SPI1, SIRT1 | 112.79 | 21.17 |
| Negative regulation by host of viral transcription | 0.02 | HDAC1, JUN | 112.79 | 21.17 |
| Cellular response to interleukin-6 | 0.02 | RELA, NFKB1 | 98.69 | 23.81 |
| Positive regulation by host of viral transcription | 0.02 | EP300, JUN | 87.73 | 26.35 |
| miRNA mediated inhibition of translation | 0.02 | ZFP36, STAT3 | 87.73 | 26.35 |
| Cellular response to interleukin-1 | 0.06 | RELA, NFKB1 | 30.37 | 58.71 |
| Negative regulation of fat cell differentiation | 0.06 | SIRT1, DDIT3 | 29.24 | 60.09 |
| Regulation of inflammatory response | 0.08 | RELA, ESR1 | 23.93 | 67.47 |

**Table: 4C** Molecular Function of Transcription factors.

| Term | PValue | Genes | Fold Enrichment | FDR |
| --- | --- | --- | --- | --- |
| RNA polymerase II core promoter proximal region sequence-specific DNA binding | 0 | HDAC2, HSF1, HDAC1, RELA, ETS2, SPI1, IRF1, ESR1, STAT3, DDIT3 | 16.43 | 0 |
| RNA polymerase II distal enhancer sequence-specific DNA binding | 0 | HDAC2, HDAC1, REL, JUN, RELA, SPI1, NFKB1 | 47.67 | 0 |
| Transcription factor activity, sequence-specific DNA binding | 0 | CEBPA, REL, XBP1, CREM, ESR1, FOXP3, AHR, STAT3, DDIT3 | 11.66 | 0 |
| Transcriptional activator activity, RNA polymerase II core promoter proximal region sequence-specific binding | 0 | RELA, SPI1, IRF1, ESR1, STAT3, KLF4, DDIT3 | 16.29 | 0 |
| Transcriptional activator activity, RNA polymerase II distal enhancer sequence-specific binding | 0 | REL, RELA, SPI1, NFKB1 | 74.91 | 0.02 |
| Core promoter binding | 0 | HDAC1, FOXP3, RUNX1, KLF4 | 37.46 | 0.14 |
| Sequence-specific DNA binding | 0 | CEBPA, XBP1, CREM, ESR1, FOXP3, DDIT3 | 9.52 | 0.3 |
| Transcriptional repressor activity, RNA polymerase II core promoter proximal region sequence-specific binding | 0 | HSF1, RELA, ETS2, SPI1 | 20.25 | 0.87 |
| DNA binding | 0 | ZFP36, EGR1, REL, ESR1, STAT1, AHR, STAT3 | 5.52 | 1.09 |
| Core promoter sequence-specific DNA binding | 0 | CREM, ESR1, SIRT1 | 43.22 | 1.98 |
| chromatin DNA binding | 0 | EP300, RELA, STAT3 | 29.57 | 4.15 |
| RNA polymerase II core promoter sequence-specific DNA binding | 0.01 | EP300, HSF1, SPI1 | 24.97 | 5.74 |
| Transcriptional activator activity, RNA polymerase II transcription regulatory region sequence-specific binding | 0.01 | NR1I2, EP300, RUNX1 | 21.2 | 7.81 |
| chromatin binding | 0.02 | HSF1, JUN, ESR1, NFKB1 | 6.17 | 21.42 |
| Histone deacetylase activity | 0.03 | HDAC2, HDAC1 | 68.1 | 24.67 |
| RNA polymerase II transcription factor activity, ligand-activated sequence-specific DNA binding | 0.05 | NR1I2, STAT3 | 37.46 | 40.27 |

**Table: 4D** KEGG pathway of Transcription Factor:

| Term | PValue | Genes | Fold Enrichment | FDR |
| --- | --- | --- | --- | --- |
| HTLV-I infection | 0 | ZFP36, EGR1, E2F1, EP300, XBP1, CREM, JUN, RELA, ETS2, SPI1, NFKBIA, NFKB1 | 11.87 | 0 |
| Hepatitis B | 0 | E2F1, EP300, JUN, RELA, NFKBIA, NFKB1, STAT1, STAT3 | 15.23 | 0 |
| Viral carcinogenesis | 0 | HDAC2, EP300, HDAC1, REL, JUN, RELA, NFKBIA, NFKB1, STAT3 | 11.19 | 0 |
| Hepatitis C | 0 | RELA, IRF1, NFKBIA, NFKB1, STAT1, STAT3 | 12.36 | 0.09 |
| Influenza A | 0 | EP300, JUN, RELA, NFKBIA, NFKB1, STAT1 | 8.9 | 0.43 |
| Toll-like receptor signaling pathway | 0 | JUN, RELA, NFKBIA, NFKB1, STAT1 | 13.14 | 0.48 |
| Herpes simplex infection | 0 | EP300, JUN, RELA, NFKBIA, NFKB1, STAT1 | 7.39 | 1 |
| Measles | 0 | RELA, NFKBIA, NFKB1, STAT1, STAT3 | 10.22 | 1.22 |
| B cell receptor signaling pathway | 0 | JUN, RELA, NFKBIA, NFKB1 | 16.73 | 1.6 |
| Chemokine signaling pathway | 0 | RELA, NFKBIA, NFKB1, STAT1, STAT3 | 8.02 | 2.95 |
| HIF-1 signaling pathway | 0 | EP300, RELA, NFKB1, STAT3 | 11.15 | 5 |
| T cell receptor signaling pathway | 0 | JUN, RELA, NFKBIA, NFKB1 | 11.04 | 5.14 |
| TNF signaling pathway | 0.01 | JUN, RELA, NFKBIA, NFKB1 | 10.04 | 6.66 |
| Cytosolic DNA-sensing pathway | 0.02 | RELA, NFKBIA, NFKB1 | 14.04 | 17.53 |
| NF-kappa B signaling pathway | 0.04 | RELA, NFKBIA, NFKB1 | 9.1 | 35.32 |
| MAPK signaling pathway | 0.06 | JUN, RELA, NFKB1, DDIT3 | 4.38 | 46.33 |
| FoxO signaling pathway | 0.08 | EP300, SIRT1, STAT3 | 6.18 | 58.67 |

**Table: 5A** Localization of MicroRNA in different cellular components.

| Subcategory | P-value | Negative LOG10(P Values) | Observed | miRNAs/precursors |
| --- | --- | --- | --- | --- |
| Microvesicle | 0 | 4.15 | 9 | hsa-miR-24-3p; hsa-miR-204-5p; hsa-miR-203a-3p; hsa-miR-106a-5p; hsa-miR-124-3p; hsa-let-7c-5p; hsa-miR-1-3p; hsa-miR-98-5p; hsa-miR-155-5p |
| Nucleus | 0 | 4.14 | 9 | hsa-miR-24-3p; hsa-miR-204-5p; hsa-miR-335-5p; hsa-miR-203a-3p; hsa-miR-106a-5p; hsa-let-7c-5p; hsa-miR-1-3p; hsa-miR-98-5p; hsa-miR-155-5p |
| exosome | 0 | 4.1 | 8 | hsa-miR-24-3p; hsa-miR-204-5p; hsa-miR-335-5p; hsa-miR-203a-3p; hsa-miR-124-3p; hsa-let-7c-5p; hsa-miR-98-5p; hsa-miR-155-5p |
| Circulating | 0 | 3.92 | 10 | hsa-miR-24-3p; hsa-miR-204-5p; hsa-miR-335-5p; hsa-miR-203a-3p; hsa-miR-106a-5p; hsa-miR-124-3p; hsa-let-7c-5p; hsa-miR-1-3p; hsa-miR-98-5p; hsa-miR-155-5p |
| Cytoplasm | 0 | 3.6 | 7 | hsa-miR-24-3p; hsa-miR-335-5p; hsa-miR-203a-3p; hsa-miR-106a-5p; hsa-let-7c-5p; hsa-miR-1-3p; hsa-miR-98-5p |
| Mitochondrion | 0.01 | 2.29 | 7 | hsa-miR-24-3p; hsa-miR-106a-5p; hsa-miR-124-3p; hsa-let-7c-5p; hsa-miR-1-3p; hsa-miR-98-5p; hsa-miR-155-5p |
| Exosome | 0.02 | 1.62 | 10 | hsa-miR-24-3p; hsa-miR-204-5p; hsa-miR-335-5p; hsa-miR-203a-3p; hsa-miR-106a-5p; hsa-miR-124-3p; hsa-let-7c-5p; hsa-miR-1-3p; hsa-miR-98-5p; hsa-miR-155-5p |
| Microvesicle | 0.03 | 1.56 | 10 | hsa-miR-24-3p; hsa-miR-204-5p; hsa-miR-335-5p; hsa-miR-203a-3p; hsa-miR-106a-5p; hsa-miR-124-3p; hsa-let-7c-5p; hsa-miR-1-3p; hsa-miR-98-5p; hsa-miR-155-5p |

**Table: 5B** Involvement of MicroRNA in different disease.

| Subcategory | P-value | Negative log10(P Values) | miRNAs/precursors |
| --- | --- | --- | --- |
| Lymphoma, T-Cell | 0 | 5.8 | hsa-miR-24-3p; hsa-miR-203a-3p; hsa-miR-124-3p; hsa-miR-1-3p; hsa-miR-155-5p |
| Inflammatory bowel disease | 0 | 5.47 | hsa-miR-24-3p; hsa-miR-203a-3p; hsa-miR-124-3p; hsa-miR-1-3p; hsa-miR-155-5p |
| hepatitis B | 0 | 4.63 | hsa-miR-203a-3p; hsa-miR-106a-5p; hsa-miR-124-3p; hsa-miR-1-3p; hsa-miR-155-5p |
| Sepsis | 0 | 4.06 | hsa-miR-203a-3p; hsa-miR-106a-5p; hsa-miR-1-3p; hsa-miR-155-5p |
| Inflammation | 0 | 4.01 | hsa-miR-203a-3p; hsa-miR-124-3p; hsa-miR-1-3p; hsa-miR-155-5p |
| HIV Infections | 0 | 3.79 | hsa-miR-203a-3p; hsa-miR-124-3p; hsa-miR-1-3p; hsa-miR-155-5p |
| Adenoviridae Infections | 0 | 3.69 | hsa-miR-203a-3p; hsa-miR-124-3p; hsa-miR-1-3p; hsa-miR-155-5p |
| Hepatitis | 0 | 3.55 | hsa-miR-203a-3p; hsa-miR-124-3p; hsa-miR-1-3p; hsa-miR-155-5p |
| Aortic valve disease | 0 | 3.34 | hsa-miR-204-5p; hsa-miR-106a-5p |
| Hepatitis C | 0 | 3.3 | hsa-miR-203a-3p; hsa-miR-124-3p; hsa-miR-1-3p; hsa-miR-155-5p |
| Asthma | 0 | 3.29 | hsa-miR-203a-3p; hsa-miR-124-3p; hsa-miR-1-3p; hsa-miR-155-5p |
| Acute Lung Injury | 0 | 2.97 | hsa-miR-124-3p; hsa-miR-1-3p; hsa-miR-155-5p |
| HIV-1 | 0 | 2.97 | hsa-miR-124-3p; hsa-miR-1-3p; hsa-miR-155-5p |
| SARS Virus | 0 | 2.92 | hsa-miR-124-3p; hsa-miR-1-3p; hsa-miR-155-5p |
| Acute Kidney Failure | 0 | 2.99 | hsa-miR-124-3p; hsa-miR-1-3p; hsa-miR-155-5p |
| Prion disease | 0 | 2.94 | hsa-miR-124-3p; hsa-miR-1-3p; hsa-miR-155-5p |
| Infection | 0 | 2.91 | hsa-miR-24-3p; hsa-miR-124-3p; hsa-miR-1-3p; hsa-miR-155-5p |
| Viral hepatitis | 0 | 2.87 | hsa-miR-203a-3p; hsa-miR-1-3p; hsa-miR-155-5p |
| Lung disease | 0 | 2.68 | hsa-miR-24-3p; hsa-miR-204-5p; hsa-miR-124-3p; hsa-miR-1-3p; hsa-miR-98-5p; hsa-miR-155-5p |
| Chronic obstructive pulmonary disease | 0 | 2.57 | hsa-miR-24-3p; hsa-miR-203a-3p; hsa-miR-155-5p |
| Human Influenza | 0.02 | 1.75 | hsa-miR-1-3p; hsa-miR-155-5p |

**Table 5C:** Involvement of MicroRNA in Gene ontology.

| Subcategory | P-value | Negative (P Value) | miRNAs/precursors |
| --- | --- | --- | --- |
| Macrophage Chemotaxis | 0 | 8.47 | hsa-miR-24-3p; hsa-miR-335-5p; hsa-miR-124-3p; hsa-miR-1-3p; hsa-miR-98-5p; hsa-miR-155-5p |
| Positive Regulation Of B Cell Activation | 0 | 8.47 | hsa-miR-24-3p; hsa-miR-335-5p; hsa-miR-124-3p; hsa-miR-1-3p; hsa-miR-98-5p; hsa-miR-155-5p |
| Regulation Of Inflammatory Response | 0 | 7.8 | hsa-miR-24-3p; hsa-miR-204-5p; hsa-miR-335-5p; hsa-miR-203a-3p; hsa-miR-106a-5p; hsa-miR-124-3p; hsa-let-7c-5p; hsa-miR-98-5p; hsa-miR-155-5p |
| Inflammatory Response | 0 | 6.21 | hsa-miR-24-3p; hsa-miR-204-5p; hsa-miR-335-5p; hsa-miR-203a-3p; hsa-miR-106a-5p; hsa-miR-124-3p; hsa-let-7c-5p; hsa-miR-1-3p; hsa-miR-98-5p; hsa-miR-155-5p |
| Cytokine Binding | 0 | 5.9 | hsa-miR-24-3p; hsa-miR-335-5p; hsa-miR-124-3p; hsa-miR-1-3p; hsa-miR-98-5p; hsa-miR-155-5p |
| Interleukin 6 Receptor Binding | 0 | 5.46 | hsa-miR-335-5p; hsa-miR-124-3p; hsa-miR-1-3p; hsa-miR-98-5p; hsa-miR-155-5p |
| Interleukin 6 Mediated Signaling Pathway | 0 | 4.52 | hsa-miR-335-5p; hsa-miR-124-3p; hsa-miR-1-3p; hsa-miR-98-5p; hsa-miR-155-5p |
| T Cell Differentiation In Thymus | 0 | 4.5 | hsa-miR-24-3p; hsa-miR-204-5p; hsa-miR-335-5p; hsa-miR-124-3p; hsa-let-7c-5p; hsa-miR-1-3p; hsa-miR-98-5p; hsa-miR-155-5p |
| Interleukin 1 Binding | 0 | 4.16 | hsa-miR-335-5p; hsa-miR-98-5p; hsa-miR-155-5p |
| Response To Interleukin 1 | 0 | 4.14 | hsa-miR-335-5p; hsa-miR-203a-3p; hsa-miR-124-3p; hsa-miR-1-3p; hsa-miR-98-5p; hsa-miR-155-5p |
| T Cell Differentiation | 0 | 3.35 | hsa-miR-204-5p; hsa-miR-335-5p; hsa-miR-124-3p; hsa-miR-1-3p; hsa-miR-98-5p |
| Cellular Response To Interleukin 4 | 0 | 3.26 | hsa-miR-335-5p; hsa-miR-124-3p; hsa-miR-1-3p |
| Regulation Of T Cell Proliferation | 0 | 3.14 | hsa-miR-335-5p; hsa-miR-124-3p; hsa-miR-155-5p |
| Viral Entry Into Host Cell Via Membrane Fusion With The Plasma Membrane | 0 | 3.11 | hsa-miR-335-5p; hsa-miR-124-3p |
| Positive Regulation Of Interleukin 6 Biosynthetic Process | 0 | 3.03 | hsa-miR-204-5p; hsa-miR-335-5p; hsa-miR-124-3p |
| Positive Regulation Of T Cell Chemotaxis | 0 | 2.93 | hsa-miR-335-5p; hsa-miR-98-5p; hsa-miR-155-5p |
| Positive Regulation Of Interleukin 10 Production | 0 | 2.39 | hsa-miR-24-3p; hsa-miR-335-5p; hsa-miR-1-3p |
| Viral Reproduction | 0.01 | 2.23 | hsa-miR-24-3p; hsa-miR-204-5p; hsa-miR-335-5p; hsa-miR-124-3p; hsa-let-7c-5p; hsa-miR-1-3p; hsa-miR-98-5p; hsa-miR-155-5p |
| Positive Regulation Of Viral Genome Replication | 0.01 | 2.03 | hsa-miR-1-3p; hsa-miR-98-5p; hsa-miR-155-5p |
| Egress Of Virus Within Host Cell | 0.03 | 1.51 | hsa-miR-24-3p; hsa-miR-124-3p; hsa-let-7c-5p |
| Positive Regulation Of Mast Cell Chemotaxis | 0.04 | 1.44 | hsa-miR-335-5p; hsa-miR-106a-5p; hsa-miR-98-5p |
| Viral Entry Into Host Cell | 0.04 | 1.42 | hsa-miR-124-3p; hsa-miR-155-5p |

**Table: 5D** Involvement of MicroRNA in KEGG Pathways.

| Subcategory | Enrichment | P-value | Negative log p Value | Observed | miRNAs/precursors |
| --- | --- | --- | --- | --- | --- |
| Graft-versus-host disease | over-represented | 0 | 8.16 | 10 | hsa-miR-24-3p; hsa-miR-204-5p; hsa-miR-335-5p; hsa-miR-203a-3p; hsa-miR-106a-5p; hsa-miR-124-3p; hsa-let-7c-5p; hsa-miR-1-3p; hsa-miR-98-5p; hsa-miR-155-5p |
| Asthma | over-represented | 0 | 7.46 | 8 | hsa-miR-24-3p; hsa-miR-204-5p; hsa-miR-335-5p; hsa-miR-203a-3p; hsa-miR-106a-5p; hsa-let-7c-5p; hsa-miR-98-5p; hsa-miR-155-5p |
| Intestinal immune network for IgA production | over-represented | 0 | 6.78 | 10 | hsa-miR-24-3p; hsa-miR-204-5p; hsa-miR-335-5p; hsa-miR-203a-3p; hsa-miR-106a-5p; hsa-miR-124-3p; hsa-let-7c-5p; hsa-miR-1-3p; hsa-miR-98-5p; hsa-miR-155-5p |
| Inflammatory bowel disease IBD | over-represented | 0 | 5.88 | 10 | hsa-miR-24-3p; hsa-miR-204-5p; hsa-miR-335-5p; hsa-miR-203a-3p; hsa-miR-106a-5p; hsa-miR-124-3p; hsa-let-7c-5p; hsa-miR-1-3p; hsa-miR-98-5p; hsa-miR-155-5p |
| Viral protein interaction with cytokine and cytokine receptor | over-represented | 0 | 4.82 | 10 | hsa-miR-24-3p; hsa-miR-204-5p; hsa-miR-335-5p; hsa-miR-203a-3p; hsa-miR-106a-5p; hsa-miR-124-3p; hsa-let-7c-5p; hsa-miR-1-3p; hsa-miR-98-5p; hsa-miR-155-5p |
| Viral myocarditis | over-represented | 0 | 3.87 | 10 | hsa-miR-24-3p; hsa-miR-204-5p; hsa-miR-335-5p; hsa-miR-203a-3p; hsa-miR-106a-5p; hsa-miR-124-3p; hsa-let-7c-5p; hsa-miR-1-3p; hsa-miR-98-5p; hsa-miR-155-5p |
| Toll-like receptor signaling pathway | over-represented | 0 | 3.33 | 10 | hsa-miR-24-3p; hsa-miR-204-5p; hsa-miR-335-5p; hsa-miR-203a-3p; hsa-miR-106a-5p; hsa-miR-124-3p; hsa-let-7c-5p; hsa-miR-1-3p; hsa-miR-98-5p; hsa-miR-155-5p |
| IL-17 signaling pathway | over-represented | 0 | 3.17 | 10 | hsa-miR-24-3p; hsa-miR-204-5p; hsa-miR-335-5p; hsa-miR-203a-3p; hsa-miR-106a-5p; hsa-miR-124-3p; hsa-let-7c-5p; hsa-miR-1-3p; hsa-miR-98-5p; hsa-miR-155-5p |
| B cell receptor signaling pathway | over-represented | 0 | 2.96 | 10 | hsa-miR-24-3p; hsa-miR-204-5p; hsa-miR-335-5p; hsa-miR-203a-3p; hsa-miR-106a-5p; hsa-miR-124-3p; hsa-let-7c-5p; hsa-miR-1-3p; hsa-miR-98-5p; hsa-miR-155-5p |
| Th17 cell differentiation | over-represented | 0 | 2.7 | 10 | hsa-miR-24-3p; hsa-miR-204-5p; hsa-miR-335-5p; hsa-miR-203a-3p; hsa-miR-106a-5p; hsa-miR-124-3p; hsa-let-7c-5p; hsa-miR-1-3p; hsa-miR-98-5p; hsa-miR-155-5p |
| Natural killer cell mediated cytotoxicity | over-represented | 0 | 2.52 | 10 | hsa-miR-24-3p; hsa-miR-204-5p; hsa-miR-335-5p; hsa-miR-203a-3p; hsa-miR-106a-5p; hsa-miR-124-3p; hsa-let-7c-5p; hsa-miR-1-3p; hsa-miR-98-5p; hsa-miR-155-5p |
| Butanoate metabolism | over-represented | 0 | 2.45 | 5 | hsa-miR-24-3p; hsa-miR-335-5p; hsa-miR-124-3p; hsa-miR-1-3p; hsa-miR-155-5p |
| Longevity regulating pathway | over-represented | 0 | 2.37 | 10 | hsa-miR-24-3p; hsa-miR-204-5p; hsa-miR-335-5p; hsa-miR-203a-3p; hsa-miR-106a-5p; hsa-miR-124-3p; hsa-let-7c-5p; hsa-miR-1-3p; hsa-miR-98-5p; hsa-miR-155-5p |
| T cell receptor signaling pathway | over-represented | 0 | 2.34 | 10 | hsa-miR-24-3p; hsa-miR-204-5p; hsa-miR-335-5p; hsa-miR-203a-3p; hsa-miR-106a-5p; hsa-miR-124-3p; hsa-let-7c-5p; hsa-miR-1-3p; hsa-miR-98-5p; hsa-miR-155-5p |
| Cytokine-cytokine receptor interaction | over-represented | 0.01 | 2 | 10 | hsa-miR-24-3p; hsa-miR-204-5p; hsa-miR-335-5p; hsa-miR-203a-3p; hsa-miR-106a-5p; hsa-miR-124-3p; hsa-let-7c-5p; hsa-miR-1-3p; hsa-miR-98-5p; hsa-miR-155-5p |
| Influenza A | over-represented | 0.01 | 1.94 | 10 | hsa-miR-24-3p; hsa-miR-204-5p; hsa-miR-335-5p; hsa-miR-203a-3p; hsa-miR-106a-5p; hsa-miR-124-3p; hsa-let-7c-5p; hsa-miR-1-3p; hsa-miR-98-5p; hsa-miR-155-5p |
| JAK-STAT signaling pathway | over-represented | 0.02 | 1.74 | 10 | hsa-miR-24-3p; hsa-miR-204-5p; hsa-miR-335-5p; hsa-miR-203a-3p; hsa-miR-106a-5p; hsa-miR-124-3p; hsa-let-7c-5p; hsa-miR-1-3p; hsa-miR-98-5p; hsa-miR-155-5p |
| Hepatitis B | over-represented | 0.04 | 1.38 | 10 | hsa-miR-24-3p; hsa-miR-204-5p; hsa-miR-335-5p; hsa-miR-203a-3p; hsa-miR-106a-5p; hsa-miR-124-3p; hsa-let-7c-5p; hsa-miR-1-3p; hsa-miR-98-5p; hsa-miR-155-5p |
| Hepatitis C | over-represented | 0.04 | 1.36 | 10 | hsa-miR-24-3p; hsa-miR-204-5p; hsa-miR-335-5p; hsa-miR-203a-3p; hsa-miR-106a-5p; hsa-miR-124-3p; hsa-let-7c-5p; hsa-miR-1-3p; hsa-miR-98-5p; hsa-miR-155-5p |
